# Environmental Drivers and Spatial Patterns of Lassa Fever Cases in Nigeria: A GIS-Based Approach to Dynamic Susceptibility Mapping

**DOI:** 10.64898/2026.09.16.26361173

**Authors:** Argyro Panagiota Nikolouzou, Frideriki Vakkalou, Iosif Polenakis, Marios Anagnostou

## Abstract

Lassa fever is a major yet persistently neglected viral hemorrhagic disease in West Africa, with an estimated 100,000–300,000 infections and approximately 5,000 deaths reported annually across the region. Case-fatality rates are typically above 15% among hospitalized patients and may approach 50% during epidemic periods, with Nigeria accounting for the largest reported burden of confirmed cases. This study presents a pilot multi-layer GIS framework for the spatial recording, visualization, and analysis of hemorrhagic fever infections in Nigeria, focusing primarily on Lassa fever. The framework integrates epidemiological observations with environmental information to move beyond conventional static disease maps toward dynamic assessment of disease susceptibility. Confirmed Lassa fever cases recorded between 2023 and 2025 were combined with weather variables obtained from Open-Meteo, CHIRPS precipitation data, land-use information, lithological characteristics, and elevation datasets. The datasets were temporally aggregated and statistically analyzed, while multiple GIS layers were used to identify and visualize recurring spatial patterns. Ondo, Edo, Bauchi, and Taraba consistently emerged as prominent hotspot states during the study period. Weekly temporal aggregation was adopted to correspond with the reporting frequency of official epidemic surveillance, enabling short-term environmental conditions to be examined in relation to confirmed cases. The findings indicate a pronounced seasonal pattern, with increased Lassa fever activity occurring predominantly during dry periods characterized by minimal rainfall, fewer precipitation hours, and reduced humidity, particularly between December and March. Of the 28 environmental variables examined, precipitation-and humidity-related indicators demonstrated the strongest inverse associations with weekly confirmed cases, with Spearman correlation coefficients reaching *ρ* = *−*0.75. Air temperature showed the strongest and most consistent positive association, reaching *ρ* = 0.633. These findings demonstrate the potential of integrating epidemiological surveillance and environmental GIS layers to characterize spatial and temporal patterns of Lassa fever and provide a foundation for more dynamic susceptibility mapping and environmental risk assessment in Nigeria.

## 1 Introduction

Viral hemorrhagic fevers (VHFs) remain among the most demanding public health threats because of their zoonotic origin, severe clinical outcomes, epidemic potential, and close dependence on local environmental conditions [1, 2, 3, 4]. In West Africa, Ebola virus disease and Lassa fever represent two of the most important VHFs, both associated with recurrent outbreaks, social disruption, and substantial pressure on health care systems. These impacts are particularly severe in settings where laboratory capacity, public awareness, and timely access to treatment remain limited [5, 6]. For this reason, the study of hemorrhagic fever risk requires an integrated perspective that goes beyond biomedical factors and incorporates geography, environment, exposure, and vulnerability.

Modern risk assessment increasingly depends on the ability to combine heterogeneous and multi-level information, including geospatial layers, remote-sensing products, field observations, climate records, and socio-environmental indicators. In this context, the present study uses Lassa fever as an environmental health risk case study to develop and demonstrate a multi-layer GIS-based analytical framework. A persistent challenge in West Africa is that information on hemorrhagic fevers is often fragmented, highly technical, or not easily accessible to the general public. Communities need clear and practical information about animal reservoirs, transmission pathways, warning symptoms, prevention practices, and the importance of avoiding unsafe traditional practices when infection is suspected. At the same time, public health authorities and researchers require spatially explicit tools capable of identifying when and where risk is likely to increase, so that prevention and preparedness actions can be better targeted.

This study therefore aims to design a multi-layer GIS-oriented workflow for recording, visualizing, and interpreting Lassa fever risk in Nigeria during the period 2023–2025. The framework integrates weekly surveillance data with weather indicators, CHIRPS precipitation data, land-use information, lithological layers, and elevation data [7, 8, 9, 10, 11, 12, 13]. The central assumption is that Lassa fever outbreaks are not randomly distributed in either time or space. Instead, they are shaped by seasonal climatic conditions, land-use patterns, and persistent geographic zones of elevated activity.

The analysis gives priority to weekly aggregation rather than monthly or annual summaries. This temporal scale is especially important because epidemic surveillance reports are commonly released on a weekly basis, allowing reported confirmed cases to be compared more directly with the meteorological conditions that prevail during the same period. The correlation analysis highlights a clear seasonal pattern, with confirmed cases increasing during dry weeks marked by limited rainfall, reduced precipitation duration, and lower humidity, and declining during wetter periods. Among the 28 environmental factors examined, rainfall-related variables, precipitation hours, humidity, and air temperature emerged as the most informative weather indicators for interpreting weekly case variation. In parallel, the integration of thematic maps allows the analysis to move beyond numerical summaries and examine the spatial logic of recurring hotspot states.

### 1.1 Related Work

Next, we present the related work regarding the climatic and environmental drivers of Lassa Fever and the ecological, spatial, and socio-environmental determinants.

#### 1.1.1 Climatic and Environmental Drivers of Lassa Fever

The influence of climate variables on Lassa fever transmission has been extensively investigated across West Africa and Nigeria. In [14] Redding et al. demonstrate that Lassa fever occurrence and incidence in Nigeria is shaped by a combination of climate, poverty, agriculture, and urbanisation factors, while also highlighting how heterogeneous reporting and diagnostic access contribute to the patchy distribution of observed incidence. Using spatiotemporal predictive models, they show that incorporating climatic variability reduced out-of-sample predictive error by 11% compared to a baseline model. Redding et al. in [15] further establish that at the local government authority level, rainfall, poverty, agriculture, urbanisation, and housing quality collectively drive predictable seasonal surges in cases, with their models demonstrating the potential to forecast incidence surges one to two months in advance as a basis for an early-warning system.

In [16] Zhao et al. quantify the association between the disease reproduction number *R* and local rainfall across Nigerian regions, reporting that a one-unit increase in average monthly rainfall over the preceding seven months is associated with a 0.62% rise in *R*, and documenting significant spatial heterogeneity in epidemic dynamics across regions. Complementing this, Akhmetzhanov et al. in [17] employ a mathematical model integrating human incidence data, rodent population dynamics, and climatological variation to show that the seasonal migratory dynamics of *Mastomys natalensis* play a key role in regulating the cyclical pattern of Lassa fever epidemics, with peak human exposure coinciding with the early dry season and the rodent breeding period.

Fuwape et al. in [18] apply multilinear regression and generalised additive models to weekly case data from six high-risk Nigerian states, finding that dew point temperature and soil temperature are significant predictors of incidence, that most climatic variables exhibit nonlinear relationships with disease occurrence, and that a temperature rise between 17–20 °C substantially increases infection rates. In [19] John et al. conduct a retrospective study in Kogi State, confirming a consistent dry-season peak from November to March, a statistically significant positive correlation between temperature and incidence (*r* = 0.282, *p* = 0.029), and identifying temperature thresholds above 35 °C and humidity between 60–80% as potential early-warning indicators. In [20] Nchom et al. employ multiple regression analysis across thirteen northern Nigerian states, reporting that weather variables collectively account for 70% of reported Lassa fever cases (*r*^2^ = 0.70), with relative humidity and rainfall exhibiting strong negative correlations with incidence (*r* = *−*0.7 and *r* = *−*0.5 respectively), while maximum temperature shows a weak and non-significant association. In [21] Oluwadare et al. use Pearson correlation and binary logistic regression across six endemic Nigerian states to confirm an inverse relationship between rainfall and humidity and Lassa fever cases, and demonstrate through binary regression that dry-season conditions significantly increase outbreak probability, with an epidemic peak from November to April.

#### 1.1.2 Ecological, Spatial, and Socio-Environmental Determinants

Beyond purely climatic factors, several studies have examined the ecological and socio-environmental context of Lassa fever distribution. In [22] Arotolu et al. apply the MaxEnt algorithm to model Lassa fever distribution in Ondo State, identifying population density, road networks, built-up settlement, and poverty as key human factors alongside climatic and altitudinal variables, and producing a suitability map that delineates high-risk local government areas to guide resource allocation. In [23] Cadmus et al. analyse five years of outbreak data in Ondo State using nearest-neighbour statistics and regression, documenting a spatio-temporal clustering of cases and an emerging shift from rural to urban occurrence, with ecological variables including nighttime light intensity, vegetation, and market presence showing significant ward-level correlations with case counts.

In [24] Davis et al. conduct a systematic review of environmental drivers of Lassa virus across West Africa, synthesising 70 studies and identifying seasonal precipitation, land-use change, and host reservoir expansion as the principal environmental determinants of viral circulation, while underscoring significant knowledge gaps that limit understanding of these complex relationships. Madaki et al. in [25] synthesise evidence from 57 studies within a One Health framework, finding that poor housing (adjusted odds ratio 1.94) and unsafe food storage are the most proximal socio-environmental risk factors, that lagged rainfall robustly predicts seasonal outbreaks, and that urbanisation substantially increases spillover risk (odds ratio 23.3), while greater rodent biodiversity appears associated with lower risk, suggesting a dilution effect. In [26] Izah et al. review social and ecological risk factors in Nigeria, implicating weak environmental hygiene, poor housing, indiscriminate waste disposal, inadequate food storage, deforestation, and poor agricultural practices as conditions that exacerbate Lassa fever transmission, and calling for sustainable management strategies that integrate these dimensions. In [27] Fichet-Calvet and Rogers present risk maps for Lassa fever in West Africa using discriminant analytical techniques applied to MODIS satellite-derived environmental variables, finding that rainfall is the dominant predictor and that predicted endemic zones cover approximately 80% of Sierra Leone and Liberia, 50% of Guinea, and 40% of Nigeria.

### 1.2 Motivation and Contribution

The main contribution of this paper is methodological. It demonstrates how diverse epidemiological, climatic, topographic, lithological, and land-use datasets can be collected and preprocessed with Python, statistically assessed using rank-correlation analysis, visualized through comparative charts, and ultimately interpreted and rendered as thematic maps using GIS software. By linking infection data with environmental drivers, the proposed workflow provides a transferable basis for dynamic susceptibility mapping and supports a more spatially informed approach to hemorrhagic fever surveillance in Nigeria.

### 1.3 Roadmap

Section 2 presents the disease context and the rationale for environmental analysis. Section 3 describes the data sources and analytical workflow. Section 4 reports the main temporal and spatial findings together with the thematic maps. Section 5 interprets the observed patterns in relation to seasonality, geography, dynamic vulnerability mapping, and public health planning. Finally, Section 6 summarizes the main findings and outlines future research directions.

## 2 Background

This section establishes the disease-specific context required for comprehending the empirical analysis that follows in this document. We firstly discuss viral hemorrhagic fevers (VHFs) and their zoonotic origins within a broader epidemiological context. The empirical case study outlined in this work focuses solely on Lassa fever, though this approach could be expanded to Ebola virus disease examining with similar thoroughness due to its status as the most widely documented VHF in West Africa, offering a crucial epidemiological investigation. Moreover, we discuss to a durther extent why the environmental and seasonal susceptibility-mapping approach adopted in this work is particularly well suited to a rodent-reservoir-driven, endemic disease such as Lassa fever, rather than to an epidemic, human-to-human-transmitted disease such as Ebola.

### 2.1 Zoonoses and Viral Hemorrhagic Fevers

Zoonotic diseases are health conditions caused by infections that are naturally passed from vertebrate animals to humans. Ecological and human elements affect their growth and survival, such as wildlife interactions, rodent exposure, food contamination, changes in land use, deforestation, settlement patterns, sanitation practices, and human mobility [3, 4]. In Nigeria’s particular context, these factors are particularly crucial because human populations might reside near natural reservoirs yet encounter restricted access to timely diagnosis and efficient public health communication. viral Hemmorhagic Fever (VHF) represent a diverse category of acute viral infections marked by fever, systemic inflammation, modified vascular function, and, in certain instances, hemorrhagic symptoms [1, 2]. The “hemorrhagic fever” state may be somewhat deceptive since not all patients exhibit noticeable bleeding, particularly in the initial or moderate stages of the illness. This diagnostic uncertainty is one reason these illnesses continue to be hard to identify quickly in endemic areas [3]. The viruses primarily originate from the families *Arenaviridae*, *Filoviridae*, *Flaviviridae*, and similar groups, with many being zoonotic in nature [2].

Viral hemorrhagic fevers constitute a particularly important category of infectious diseases, due to the high morbidity and mortality they exhibit, as well as their potential to cause serious epidemics. Among the most important etiological factors of these diseases, as well as those we study in this paper, are, as we have seen, Ebola virus and Lassa virus. Although both cause hemorrhagic diseases and are mainly found in West Africa, they present essential differences. Specifically, in their biological structure, epidemiology, clinical picture and treatment strategies.Understanding their similarities and differences is important both for epidemiological surveillance and for prevention, diagnosis and treatment strategies. Ebola virus belongs to the Filoviridae family and the Ebolavirus genus. It is a single-stranded, negative-sense RNA virus, non-segmented, and characterized by a filamentous morphology. There are several species of the virus, with Zaire ebolavirus considered the most dangerous to humans. The structure of the virus and its ability to inhibit the host’s innate immune response contribute decisively to the severity of the disease. In contrast, Lassa fever belongs to the Arenaviridae family and the Mammarenavirus genus. It also has negative-sense RNA, but its genome is segmented into two strands, S and L. This different molecular organization has implications for the replication mechanism and genetic diversity of the virus, which affects both the diagnostic approach and the development of vaccines.

Both viruses are zoonotic, but differ significantly in their natural hosts. For Ebola virus, fruit bats are considered potential hosts, while transmission to humans can occur through contact with infected wild animals, such as primates. Human-to-human transmission occurs mainly through direct contact with body fluids of infected individuals, which explains the rapid spread in conditions of inadequate sanitary protection. In contrast, Lassa fever has as its main source the rodent Mastomys natalensis, which can live close to humans in endemic areas. Transmission to humans occurs mainly through contact with urine or feces of infected rodents, while human-to-human transmission is less common but possible, especially in hospital environments without adequate safety measures. On the one hand, Ebola virus disease occurs mainly in epidemic outbreaks in Central and West Africa. These outbreaks are characterized by high mortality rates and significant socioeconomic impacts. The largest recorded outbreak occurred in the period 2013–2016, causing international concern and mobilization of health organizations. On the other hand, Lassa virus infection is endemic in many countries in West Africa, mainly in Nigeria, and is estimated to cause hundreds of thousands of infections annually. However, a large proportion of cases remain undiagnosed, as the disease often presents with mild or non-specific symptoms.

### 2.2 Lassa Fever

Lassa fever is a serious and complex viral infection caused by Lassa virus, an RNA virus of the Arenaviridae family. It belongs to the broader group of viral hemorrhagic fevers, which are characterized by acute onset, systemic involvement and in severe cases hemorrhagic manifestations, organ dysfunction and high mortality in severely ill patients. Lassa fever was first reported and isolated in the early 1960s in Nigeria, in the small town of Lassa, from which it takes its name, and has since been considered an endemic disease for areas of West Africa. Unlike some other hemorrhagic diseases, such as Ebola, Lassa fever is not usually considered a pandemic but remains an endemic zoonosis in areas where infected rodents, the natural source of the virus, are found. The epidemiology of Lassa fever features ”silent” transmission and numerous mild or clinical cases, complicating diagnosis and monitoring of the disease [28].

Lassa fever is a single-stranded RNA virus that is primarily transmitted to humans through contact with the feces of infected rodents, primarily rats of the species Mastomys natalensis. These rodents are abundant in rural and semi-urban areas of West Africa, where they often enter homes or food storage areas, contaminating environments with their urine and feces [29]. Exposure to these contaminated environments, consumption of contaminated food, or even inhalation of aerosols containing the virus can lead to human infection. In addition, human-to-human transmission has been documented. This has primarily occurred through direct contact with blood, urine, saliva, or other contaminated body fluids, particularly in healthcare settings where preventive measures may be inadequate. However, human-to-human transmission is not common, due to the low content of the virus in the fluids.

The incubation period for Lassa fever is usually two to twenty-one days, with most cases developing symptoms within the first three weeks after exposure. Many cases of the disease are asymptomatic or have mild symptoms, leading to an underestimation of the disease’s spread in endemic areas. For symptomatic individuals, the first symptoms are general and nonspecific, including fever, weakness, malaise, headache, and myalgia. These symptoms can easily be confused with other common infections in the area, such as malaria, typhoid fever, or other viral hemorrhagic fevers. In advanced or severe clinical stages, nausea, vomiting, diarrhea, cough, chest pain, as well as hemorrhagic manifestations from mucous membranes, such as nosebleeds or gums, low blood pressure, and even seizures and shock are observed.

## 3 Materials and Methods

This section describes the data sources, preprocessing steps and analytical procedures underlying the study. We primarilly discuss the collection and storage of Lassa fever surveillance data as well as the acquisition and weekly aggregation of meteorological indicators. Next, we present the combination of these data in order to investigate seasonal susceptibility patterns and to quantify environmental associations through Spearman rank correlation, proceeding to the cartographic layers and classification methods used to represent the spatial dimension of the analysis.

### 3.1 Epidemiological Data

Organizations as the World Health Organization (WHO) and the Centers for Disease Control and Prevention (CDC) provide confirmed data that is also used in the scientific community. However, each disease, depending on the country in which it appears, may also have official government sources. Often, these sources offer additional details, including the count of cases and the specific cities or regions where they were noted, aiding in more in-depth analysis. Furthermore, scientific articles are valuable as they offer analyzed data, statistical evaluations, and findings that aid in research. In particular, Nigeria, has invested in recording and monitoring various diseases, establishing several case recording centers. This has been of great help, as the official website of the Nigeria Center for Disease Control and Prevention (NCDC) provides weekly updates on confirmed cases of Lassa virus.

This study utilized data from 2023 to 2025, verified, where possible, with the above-mentioned official international health organizations. At the same time, the areas where the cases were recorded were identified for each year from the same official source. Nigeria is a federal state divided into 36 states and has a federal capital. In turn, the states are divided into local administrative areas (LGAs – Local Government Areas) with the number reaching 774 LGAs in the entire country. According to the outbreak statistics, 30 LGAs are the most frequent areas.

In the proposed approach, the data were organized and processed as raw data where each line corresponds to a weekly record that includes information regarding the starting date of the week, ending date of the week, the year, week number, suspected cases, recorded cases and the deaths. In a similar way, there were organized data refering to cases by state and by local government area. Table 2 presents the cases on an annual basis for each state (note that, “–” indicates no LGA in that state met this threshold in that year), while Table 3 presents the cases on an annual basis by local government area (note that, “–” indicates the LGA did not report *>*10 confirmed cases in that year).

**Table 1:** Local Government areas.

| State | Local Government Areas |
| --- | --- |
| Ondo | Owo, Akure South, Akoko South West, Ose, Akure North |
| Bauchi | Kirfi, Toro, Bauchi, Tafawa Balewa, Ganjuwa |
| Edo | Etsako West, Esan West, Esan North East, Owan West, Etsako Central, Esan Central, Ovia North East, Esan South East |
| Taraba | Jalingo, Bali, Ardo Kola, Gassol |
| Benue | Makurdi |
| Ebonyi | Abakaliki, Izzi, Ikwo |
| Kogi | Idah |
| Enugu | Uzo Uwani |
| Plateau | Jos North |
| Delta | Oshimili South |

**Table 2:** Confirmed cases by state, aggregated across LGAs with *>*10 cases per year.

| State | 2023 | 2024 | 2025 |
| --- | --- | --- | --- |
| Ondo | 349 | 340 | 177 |
| Bauchi | 103 | 215 | 149 |
| Edo | 250 | 234 | 85 |
| Taraba | 59 | 79 | 63 |
| Ebonyi | 25 | 19 | – |
| Benue | 33 | 34 | – |
| Kogi | 10 | 12 | – |
| Enugu | – | 11 | – |

**Table 3:** Confirmed cases by local government area, aligned across years.

| State | LGA | 2023 | 2024 | 2025 |
| --- | --- | --- | --- | --- |
| Ondo | Owo | 224 | 180 | 76 |
| Ondo | Akure South | 59 | 72 | 53 |
| Ondo | Akoko South West | 16 | 38 | 33 |
| Ondo | Ose | 13 | 34 | 15 |
| Ondo | Akure North | 37 | 16 | – |
| Bauchi | Kirfi | 48 | 62 | 62 |
| Bauchi | Toro | 32 | 102 | 52 |
| Bauchi | Bauchi | 23 | 29 | 23 |
| Bauchi | Tafawa Balewa | – | 11 | 12 |
| Bauchi | Ganjuwa | – | 11 | – |
| Edo | Etsako West | 118 | 103 | 55 |
| Edo | Esan West | 61 | 63 | 15 |
| Edo | Esan North East | 48 | 44 | 15 |
| Edo | Owan West | 12 | 12 | – |
| Edo | Etsako Central | – | 12 | – |
| Edo | Esan Central | 11 | – | – |
| Taraba | Jalingo | 35 | 24 | 28 |
| Taraba | Bali | 24 | 25 | 24 |
| Taraba | Ardo Kola | – | 19 | 11 |
| Taraba | Gassol | – | 11 | – |
| Ebonyi | Abakaliki | 25 | 19 | – |
| Benue | Makurdi | 33 | 34 | – |
| Kogi | Idah | 10 | 12 | – |
| Enugu | Uzo Uwani | – | 11 | – |

### 3.2 Meteorological Data

Weather data spanning from 2023 to 2025 were retrieved through Open Meteo [9], a platform that collects information from various meteorological stations and provides it for free. By comparing, selecting coordinates, the longitude and latitude of an area, the time period and the weather parameters of interest for the analysis, a file is created. For each of the 30 local government areas studied, a separate file was created, where the coordinates used for each area are those of the largest city. In the generated files, each line corresponds to the values of the parameters requested on a daily basis. Table 4 presents all weather parameters utilized in this study.

**Table 4:**
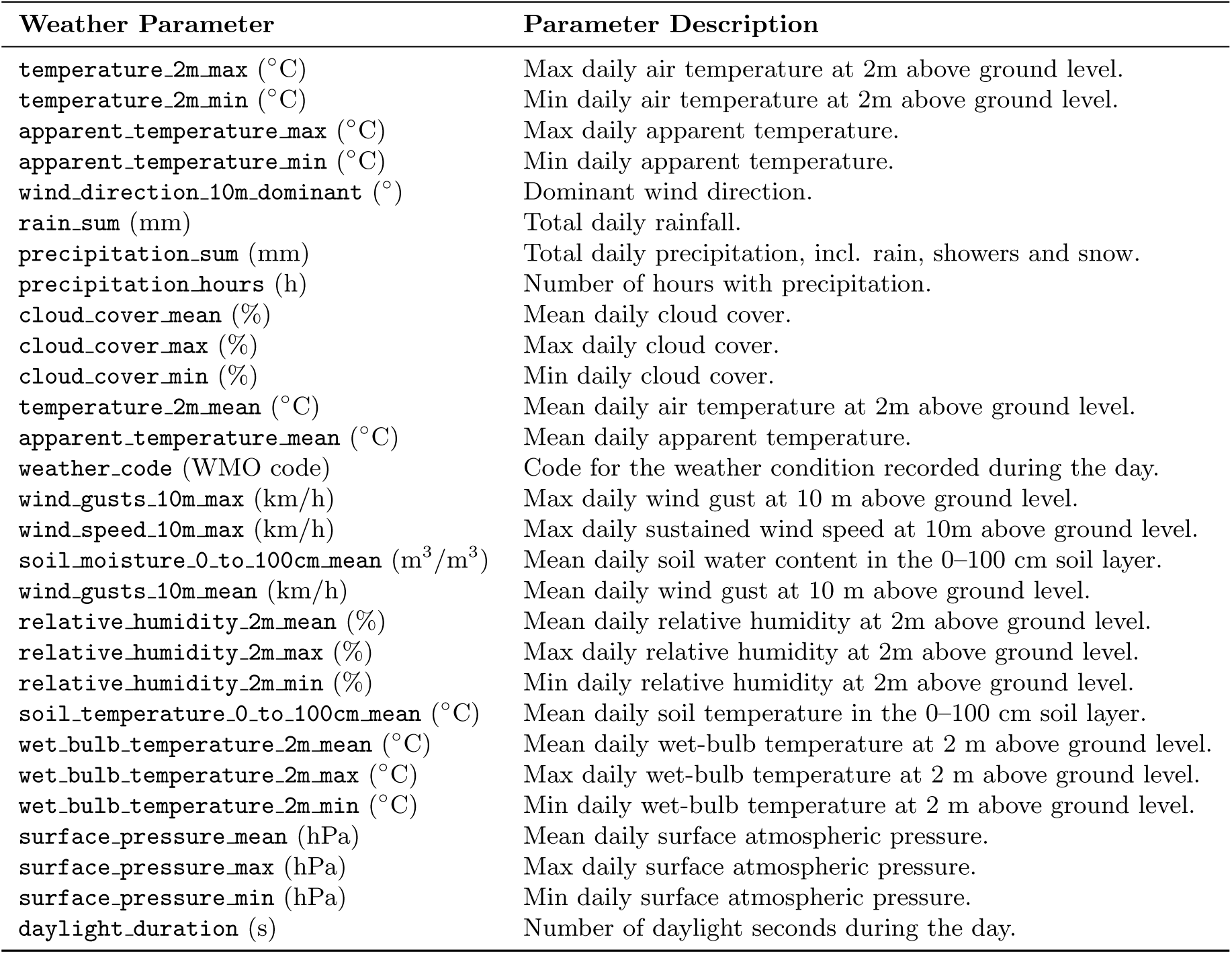
Main weather variables used in the Lassa fever seasonality analysis.

| Weather Parameter | Parameter Description |
| --- | --- |
| temperature_2m_max (°C) | Max daily air temperature at 2m above ground level. |
| temperature_2m_min (°C) | Min daily air temperature at 2m above ground level. |
| apparent_temperature_max (°C) | Max daily apparent temperature. |
| apparent_temperature_min (°C) | Min daily apparent temperature. |
| wind_direction_10m_dominant (°) | Dominant wind direction. |
| rain_sum (mm) | Total daily rainfall. |
| precipitation_sum (mm) | Total daily precipitation, incl. rain, showers and snow. |
| precipitation_hours (h) | Number of hours with precipitation. |
| cloud_cover_mean (%) | Mean daily cloud cover. |
| cloud_cover_max (%) | Max daily cloud cover. |
| cloud_cover_min (%) | Min daily cloud cover. |
| temperature_2m_mean (°C) | Mean daily air temperature at 2m above ground level. |
| apparent_temperature_mean (°C) | Mean daily apparent temperature. |
| weather_code (WMO code) | Code for the weather condition recorded during the day. |
| wind_gusts_10m_max (km/h) | Max daily wind gust at 10 m above ground level. |
| wind_speed_10m_max (km/h) | Max daily sustained wind speed at 10m above ground level. |
| soil_moisture_0_to_100cm_mean (m<sup>3</sup>/m<sup>3</sup>) | Mean daily soil water content in the 0–100 cm soil layer. |
| wind_gusts_10m_mean (km/h) | Mean daily wind gust at 10 m above ground level. |
| relative_humidity_2m_mean (%) | Mean daily relative humidity at 2m above ground level. |
| relative_humidity_2m_max (%) | Max daily relative humidity at 2m above ground level. |
| relative_humidity_2m_min (%) | Min daily relative humidity at 2m above ground level. |
| soil_temperature_0_to_100cm_mean (°C) | Mean daily soil temperature in the 0–100 cm soil layer. |
| wet_bulb_temperature_2m_mean (°C) | Mean daily wet-bulb temperature at 2 m above ground level. |
| wet_bulb_temperature_2m_max (°C) | Max daily wet-bulb temperature at 2 m above ground level. |
| wet_bulb_temperature_2m_min (°C) | Min daily wet-bulb temperature at 2 m above ground level. |
| surface_pressure_mean (hPa) | Mean daily surface atmospheric pressure. |
| surface_pressure_max (hPa) | Max daily surface atmospheric pressure. |
| surface_pressure_min (hPa) | Min daily surface atmospheric pressure. |
| daylight_duration (s) | Number of daylight seconds during the day. |

In our study weather data are stored on a daily basis, while infection case data are stored on a weekly basis. This difference creates an incompatibility in the data that prevents their comparison and the overall investigation. In addition, infection case data cannot be converted to a daily basis, so converting weather data to a weekly basis performed to match meteorological to epidemiological data. The corresponding procedure, developed in Python, reads the daily meteorological data file from Open Meteo, groups them based on the ISO year and the ISO week number and calculates the weekly averages for all weather parameters. Finally, 3 additional attributes are added, one for the week number, one with the starting date of the week, and one for the ending date of the week, while the final results are exported to a new Excel file.

### 3.3 Seasonality Research

This study examines the relationship between confirmed Lassa fever cases and meteorological conditions (see, Table 4). Preliminary observations suggested that lower numbers of confirmed cases tended to occur during wetter periods. To investigate this relationship systematically, weekly confirmed case counts were compared with corresponding meteorological parameters. As the epidemiological data were available at a weekly temporal resolution, the meteorological data were aggregated accordingly. For each study year, only the Local Government Areas (LGAs) reporting confirmed Lassa fever cases were included in the analysis. Weekly mean values of the selected meteorological variables were then calculated for these LGAs, allowing direct comparison between disease occurrence and environmental conditions.

To facilitate the interpretation of epidemic intensity, an operational threshold was established to identify outbreak weeks. The mean number of weekly confirmed cases was calculated separately for each year of the study period, and the highest annual mean weekly case count was selected as a reference value representing elevated transmission activity. Based on this criterion, weeks reporting more than 25 confirmed cases were classified as outbreak weeks. This threshold intended to serve as a study-specific indicator for distinguishing periods of increased Lassa fever activity from routine surveillance periods. The resulting classification was used to support comparisons between epidemic peaks and concurrent environmental conditions, particularly rainfall, precipitation duration, humidity, and temperature.

Correlation analysis was used to examine whether weekly variations in confirmed Lassa fever cases were systematically associated with changes in environmental and meteorological conditions. Since the purpose of the analysis was not to build a predictive model, but to identify the direction and relative strength of monotonic associations, a non-parametric approach was selected. This was particularly appropriate for the present dataset, where epidemiological and environmental variables may show skewed distributions, seasonal fluctuations, and non-linear patterns.

In particular, in opur study we utilzied Spearman’s rank correlation coefficient (*ρ*, or *r_s_*), a statistical measure that evaluates the strength and direction of a monotonic relationship between two variables [30]. Unlike Pearson’s correlation coefficient, which only detects linear relationships and is based on assumptions such as normality, homoscedasticity, and interval or ratio data, Spearman’s coefficient is calculated from the ranks of the values rather than the raw data. This makes it suitable for ordinal data, data that differ from a normal distribution, or for relationships that are monotonic but not purely linear [31]. In the simple case of no tied ranks, the coefficient is calculated as:

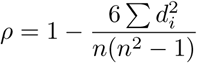

where *d_i_* signifies the difference between the positions of each pair of observations and *n* indicates the size of the sample. In practice, however, datasets frequently contain tied values, rendering the formula above just an estimation. A more precise method in these scenarios is to use the Pearson formula on the ranked data, which appropriately handles ties by giving equal ranks to tied values [32, 33].

Spearman’s coefficient requires comparatively fewer assumptions than Pearson’s: (a) the data must be at least ordinal, (b) the relationship between the variables must be monotonic (though not necessarily linear), and (c) observations must be independent of one another [34]. No assumption of normality is required, which makes the coefficient particularly useful when data exhibit substantial skewness or contain outliers. The values of *ρ* range between *−*1 and +1. Values approaching +1 indicate a strong positive monotonic correlation (as one variable increases, the other does as well), values close to *−*1 signify a strong negative correlation, and values near 0 suggest a lack of a monotonic relationship [34].Investigators frequently utilize recognized benchmarks to assess the strength of correlation, like the criteria suggested by Cohen [35] for Pearson *r* (around 0.10 = small, 0.30 = moderate, 0.50 = large effect), which are also similarly employed for Spearman’s *ρ*, although there are reservations about their overall applicability [36].

Bivariate correlations between weekly environmental/meteorological parameters and weekly confirmed cases were assessed using the Spearman rank correlation coefficient (*ρ*). The data utilized are identical to those employed to create the graphs. Prior to analysis, non-numeric values (like decimal points) were transformed into numeric format. The predictor variables for the candidate set included all numeric columns, excluding identifiers and the outcome variable (i.e., columns related to total cases, deaths, year, week, date/time, or sheet labels). This resulted in 28 environmental factors, including temperature, apparent temperature, relative humidity, precipitation, cloud cover, wind speed/gust/direction, surface pressure, soil temperature and moisture, wet bulb temperature, and daylight duration.

For every predictor, Spearman’s *ρ* was calculated in relation to the weekly confirmed case count utilizing the spearmanr function from the SciPy library in Python [37], while data manipulation was conducted using pandas [38]. For every variable pair, observations with missing values in either variable were removed on a pairwise basis (listwise deletion per variable pair), and a correlation was calculated only if at least three valid paired observations persisted and both variables showed non-zero variance. Because the outcome variable contained six weeks with missing case data, this resulted in *N* = 150 valid weekly observations for each of the 28 correlations performed. The sign of *ρ* was retained to indicate the direction of the relationship between each environmental parameter and case incidence (positive or negative).

### 3.4 Cartographic Layers

In addition to the weekly weather tables used to compare epidemic weeks, discrete climate layers were applied in a format to the mapping aspect of the study. The state-level administrative boundary shapefile of Nigeria was obtained from the Humanitarian Data Exchange (HDX) platform, where it is maintained as the Common Operational Dataset (COD) for administrative boundaries of Nigeria [39]. The shapefile was produced by the Office for the Surveyor General of the Federation of Nigeria (OSGOF) in collaboration with the United Nations Cartographic Section and eHealth Africa. The state boundary layer was incorporated into all thematic maps produced in this study, including the case distribution maps, the land-use, lithological and elevation maps, and the annual precipitation maps, as an overlaid reference layer to delineate state divisions and facilitate spatial interpretation across all cartographic outputs. For the case distribution maps specifically, annual confirmed Lassa fever case totals by state were additionally joined manually to the attribute table of this shapefile for each year of the study period (2023–2025), using the state name as the common identifier, and the resulting attribute table served as the input for the choropleth classification and cartographic rendering carried out in ArcGIS Pro. The precipitation maps were created using CHIRPS data provided through [10], is a widely used precipitation dataset that combines satellite-based infrared precipitation estimates with ground-based data, providing gridded precipitation time series, examining precipitation variability, and conducting hydroclimatic research [10]. CHIRPS precipitation data was already provided at an annual level. The country precipitation maps used the CHIRPS dataset, while the LGA weekly weather assessment used Open-Meteo data [9].

Beyond the climate and epidemiological layers, additional static contextual layers were incorporated into the cartographic analysis: elevation, lithology and land use/land cover. Elevation data were obtained from NASA Earthdata’s Digital Elevation/Terrain Model holdings [11]; all DEM tiles covering the national territory were downloaded, mosaicked into a single continuous raster, and subsequently clipped to the national boundary of Nigeria to produce the elevation layer used in Figure 10. The lithological layer was derived from the Africa Surface Lithology dataset, developed by the U.S. Geological Survey and The Nature Conservancy and distributed through the RCMRD Open Data portal [12], and was classified using the Unique Values method, given its categorical (rather than continuous) nature. The land-use/land-cover layer was obtained from the ORNL DAAC annual Land Use and Urban Land Cover product for Ethiopia, Nigeria and South Africa (2016–2020) [13], and was likewise classified using Unique Values for the reasons detailed below.

Fixed manual classification intervals were used for spatial representation of precipitation, rather than classification methods based on data distributions. This choice was made to ensure direct comparability between different years, as methods based on the same data produce different class boundaries for each map and, consequently, the same color may correspond to different ranges of values from year to year. In contrast, by using fixed boundaries, each color class represents the same precipitation range across all annual maps. Annual rainfall was classified into this five fixed classes, with limits of 800, 1,200, 1,600 and 2,000 mm. The final classes were: *≤*800 mm, *>*800–1,200 mm, *>*1,200–1,600 mm, *>*1,600–2,000 mm and *>*2,000 mm. These intervals reflect the known north–south rainfall gradient of Nigeria, from the drier Sahelian and Sudanese zones in the north to the wetter Guinean and coastal zones in the south. The use of fixed rainfall thresholds enables consistent comparison of spatial rainfall patterns between different years and allows identification of persistent wet and dry areas.

Various classification methods were employed for the cartographic depiction of epidemiological variables, based on the characteristics and distribution of the data. For the state-level case maps, a common manual classification scheme was adopted for the entire study period. Confirmed cases were divided into five classes: 0, 1–10, 11–40, 41–80 and more than 80 cases. The zero value was retained as an independent category in order to distinguish states with no reported cases from states in which at least one case was recorded. The 1–10 class represents limited occurrence, whereas the 11–40 and 41–80 classes distinguish progressively higher levels of state-level burden. The final open-ended class, exceeding 80 cases, was used to isolate the comparatively small number of states with exceptionally high case counts and to identify the principal epidemiological hotspots.

The unequal widths of the intervals were intentional and reflect the strongly right-skewed distribution of the epidemiological data, in which many states recorded zero or relatively few cases, while only a limited number exhibited substantially higher values. Narrower intervals were therefore retained at the lower end of the distribution to preserve differences among states with low case counts, whereas broader intervals were used at higher values to accommodate the increasing dispersion of the observations. The class boundaries were defined through joint examination of the annual state-level case distributions for 2023–2025 and were applied unchanged to all three maps. Consequently, a given case count is represented by the same cartographic class in every year, allowing changes in both hotspot location and burden category to be interpreted consistently. Annual maximum values were reported separately, since they describe changes in the intensity of the highest state-level burden that would not be visible from the open-ended upper class alone. These intervals should therefore be understood as cartographic and comparative thresholds developed for this study, rather than as official clinical or public-health risk categories.

Conversely, for the mapping of land uses/covers and lithology, a Unique Values classification was utilized since these data represent categorical rather than continuous numerical variables. Each raster or thematic layer value relates to a particular land cover or lithological category, including urban regions, agricultural land, shrubland, forest areas, bodies of water or coastal regions, in the case of land use, and alluvial, metasedimentary, non-carbonate or other parent-material classes, in the case of lithology. Consequently, applying continuous classification techniques like Jenks or equal intervals would be unsuitable, as it would change the thematic significance of the categories. Classification of Unique Values maintains the original categorical information and enables the direct interpretation of each class as a separate type of land cover or lithological unit.

In general, the cartographic classification approach was adapted to the measurement scale, distribution and analytical purpose of each variable. For the state-level case maps, a common manual classification scheme was applied consistently throughout the study period in order to preserve direct comparability between years and to distinguish states with no reported cases, low or intermediate burdens, and the principal epidemiological hotspots. Annual precipitation was likewise represented using fixed manual classes so that identical colours corresponded to the same rainfall ranges in every annual map, thereby supporting the comparison of persistent wet and dry spatial patterns. By contrast, land-use/land-cover and lithology were represented using the Unique Values method because their values correspond to discrete thematic categories rather than to continuous numerical measurements. The elevation layer was treated as a continuous geomorphological surface, allowing the interpretation of relief and spatial elevation gradients while retaining its quantitative character. Taken together, these choices provide a cartographic framework that is temporally consistent for the annual epidemiological and climatic layers, thematically faithful for the categorical layers, and spatially informative for the continuous elevation surface. The resulting maps therefore support a coherent, interpretable and visually reliable comparison of the epidemiological, climatic and environmental conditions examined in the study.

## 4 Results

This section presents the findings of the study in three complementary stages. First, weekly confirmed cases are compared against key weather indicators to identify recurring temporal susceptibility signals. Next, the Spearman rank-correlation analysis quantifies the strength and direction of these associations across all 28 environmental parameters examined. Finally, the spatial dimension of the analysis is addressed through a multi-layer cartographic interpretation, linking the recurring hotspot states to their land-use, lithological, elevational and precipitation context.

### 4.1 Temporal Susceptibility Signals Based on Weekly Weather Indicators

To examine to a further extent the temporal structure of Lassa fever occurrence, weekly confirmed cases were compared with four weather-related indicators for the available charted years: rainfall amount, precipitation duration, mean relative humidity and daylight duration. These indicators were selected because they showed the clearest contrast between outbreak and non-outbreak weeks during the preprocessing stage. The purpose of this visual comparison was not to establish direct causality, but to identify recurring temporal susceptibility signals that may support early-warning interpretation and dynamic risk mapping. Next, there are discussed particular meteorological drivers that affect exhibit a potential relation to the spread of the pathogen in terms of confirmed cases, namely, the rainfall ammount, the precipitation hours, the relativehumidity, and the daylight duration (note that in Figures 1—4, the red line represents weekly confirmed Lassa fever cases, while the blue line represents the corresponding weekly environmental factor derived from the processed weather dataset [9]):

- **Rainfall Amount:** During the examined period, the main increases in confirmed Lassa fever cases occur during weeks with very low or near-zero rainfall. This trend is noticeable in 2023, 2024, and 2025, with peak case values occurring in the initial epidemiological weeks, prior to the main rainy season. With the rise in rainfall in the year’s midsection, confirmed cases decrease and stay relatively low. These diagrams support the interpretation that rainfall is not necessarily an immediate direct driver of transmission, but rather a seasonal environmental indicator that structures the ecological conditions under which the rodent reservoir and human populations interact. This agrees with the broader seasonal explanation of Lassa fever transmission in Nigeria, according to which dry-season conditions are associated with increased human exposure to infected rodents [22, 40]. Figure 1 provides a clear visual evidence of the dry-season signal.
- **Precipitation Hours:** The highest numbers of confirmed cases are concentrated in weeks with few or no precipitation hours, while weeks with longer precipitation duration mainly occur during the middle part of the year, when confirmed cases are comparatively low. This is evident in 2023, 2024 and 2025, where outbreak peaks occur during early weeks with few or no precipitation hours; the subsequent increase in precipitation duration coincides with comparatively lower case numbers. The repeated inverse visual pattern between precipitation duration and confirmed cases supports the conclusion that dry-period indicators are central to the temporal susceptibility profile of Lassa fever in Nigeria. Figure 2 reinforces the signal observed in the rainfall diagrams.
- **Relative Humidity:** In 2023 and 2024, the peak outbreak in early year occurs before the prolonged high-humidity phase. In 2025, verified cases once more hit their peak numbers in the initial weeks, prior to humidity attaining its highest seasonal rates. Humidity should not be viewed as a sole causal element; instead, it serves as a supporting moisture-related signal that aids in differentiating dry-season outbreak situations from wetter non-outbreak phases. Figure 3 indicates that the primary peaks in confirmed cases typically arise prior to the time of maximum relative humidity.
- **Daylight Duration:** The primary peaks of confirmed cases happen in the early part of the year, prior to the time of longest daylight duration, and in certain years, further rises can be observed toward the year’s end as daylight duration diminishes again. This trend is apparent in 2023 and 2024, as late-year rises align with the shift back to shorter days. The duration of daylight thus aids in placing the outbreaks within the yearly seasonal cycle. Figure 4 should be interpreted primarily as evidence of seasonality rather than as evidence of a direct biological effect. Overall, the weekly diagrams indicate that Lassa fever activity is concentrated mainly during dry-season weeks. Confirmed cases are highest when rainfall and precipitation hours are very low, while the increase of rainfall, precipitation duration and relative humidity during the middle of the year coincides with a marked reduction in cases. Daylight duration further supports the seasonal interpretation by showing that the main peaks occur within a repeated annual timing window. These findings support the interpretation that Lassa fever risk follows a recurring temporal susceptibility pattern, which can be represented through multi-layer GIS analysis and can later be expanded into more formal early-warning and dynamic vulnerability-mapping approaches.

**Figure 1:**
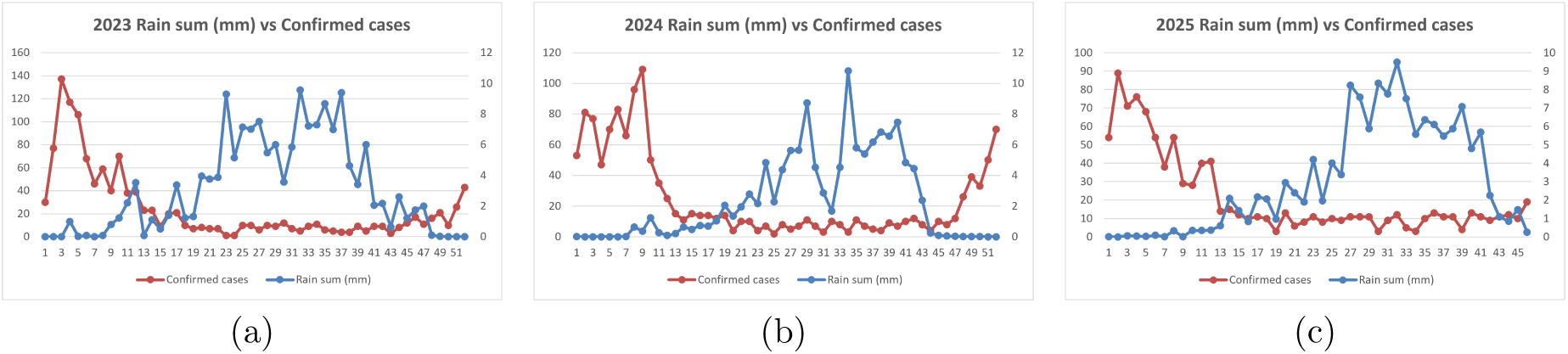
Weekly rain sum and confirmed Lassa fever cases in Nigeria for 2023–2025.

**Figure 2:**
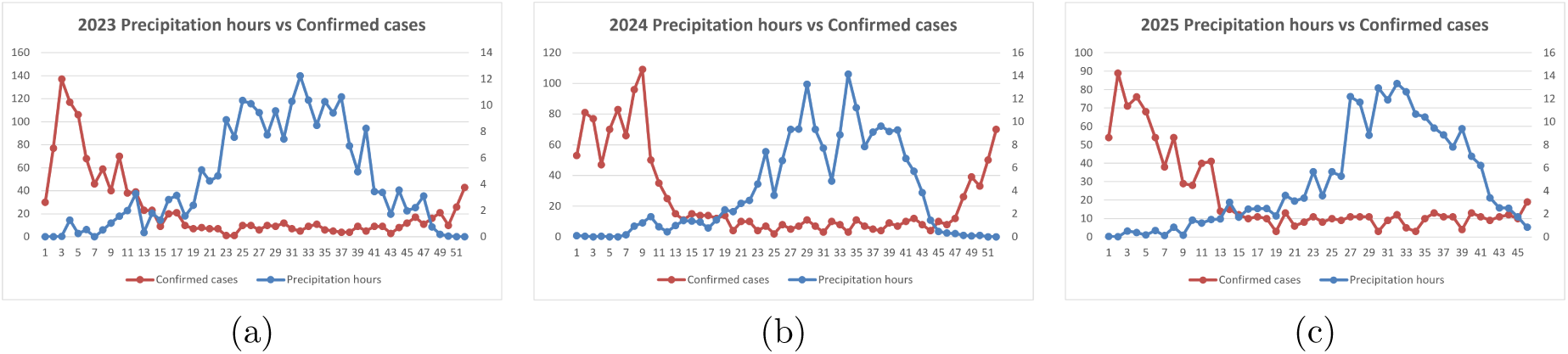
Weekly precipitation hours and confirmed Lassa fever cases in Nigeria for 2023–2025.

**Figure 3:**
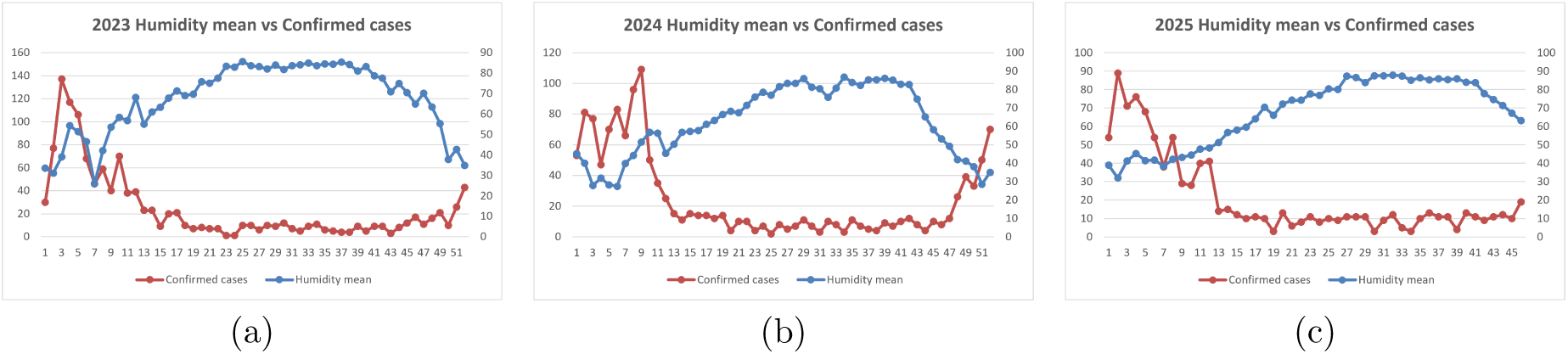
Weekly mean relative humidity and confirmed Lassa fever cases in Nigeria for 2023–2025.

**Figure 4:**
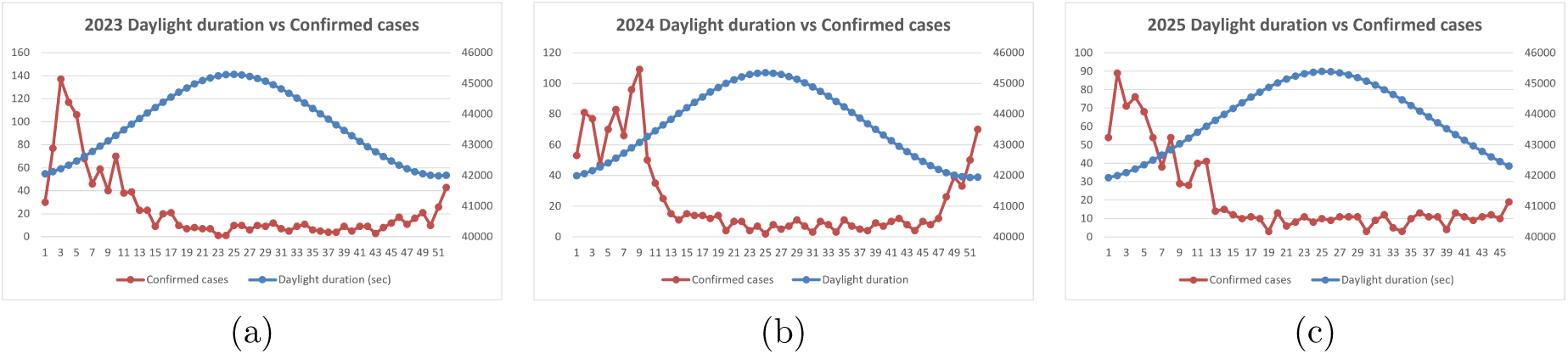
Weekly daylight duration and confirmed Lassa fever cases in Nigeria for 2023–2025.

### 4.2 Correlation Results

In Table 5 there are presented the Spearman correlation results for weekly confirmed Lassa fever cases and environmental parameters, where from the 28 environmental factors analyzed, the majority showed a non-negligible Spearman correlation with weekly confirmed Lassa fever cases; only maximum apparent temperature showed a coefficient close to zero (*ρ* = 0.033), suggesting no substantial monotonic relationship with case occurrence.

**Table 5:** Spearman correlation results for weekly confirmed Lassa fever cases and environmental parameters.

| Parameter | $N$ | $\rho$ | Strength | Direction |
| --- | --- | --- | --- | --- |
| relative_humidity_2m_min (%) | 150 | -0.752 | Very strong | Negative |
| relative_humidity_2m_mean (%) | 150 | -0.749 | Very strong | Negative |
| relative_humidity_2m_max (%) | 150 | -0.746 | Very strong | Negative |
| precipitation_hours (h) | 150 | -0.728 | Very strong | Negative |
| rain_sum (mm) | 150 | -0.703 | Very strong | Negative |
| precipitation_sum (mm) | 150 | -0.703 | Very strong | Negative |
| wet_bulb_temperature_2m_min (°C) | 150 | -0.697 | Strong | Negative |
| wet_bulb_temperature_2m_mean (°C) | 150 | -0.660 | Strong | Negative |
| temperature_2m_max (°C) | 150 | 0.633 | Strong | Positive |
| soil_moisture_0_to_100cm_mean (m<sup>3</sup>/m<sup>3</sup>) | 150 | -0.626 | Strong | Negative |
| wet_bulb_temperature_2m_max (°C) | 150 | -0.618 | Strong | Negative |
| daylight_duration (s) | 150 | -0.612 | Strong | Negative |
| cloud_cover_mean (%) | 150 | -0.592 | Strong | Negative |
| cloud_cover_min (%) | 150 | -0.587 | Strong | Negative |
| wind_direction_10m_dominant (°) | 150 | -0.562 | Strong | Negative |
| cloud_cover_max (%) | 150 | -0.523 | Strong | Negative |
| apparent_temperature_min (°C) | 150 | -0.496 | Moderate | Negative |
| temperature_2m_mean (°C) | 150 | 0.414 | Moderate | Positive |
| surface_pressure_min (hPa) | 150 | -0.311 | Moderate | Negative |
| surface_pressure_mean (hPa) | 150 | -0.304 | Moderate | Negative |
| soil_temperature_0_to_100cm_mean (°C) | 150 | 0.300 | Moderate | Positive |
| wind_speed_10m_max (km/h) | 150 | 0.289 | Weak | Positive |
| wind_gusts_10m_mean (km/h) | 150 | 0.274 | Weak | Positive |
| wind_gusts_10m_max (km/h) | 150 | 0.269 | Weak | Positive |
| apparent_temperature_mean (°C) | 150 | -0.237 | Weak | Negative |
| temperature_2m_min (°C) | 150 | -0.225 | Weak | Negative |
| surface_pressure_max (hPa) | 150 | -0.225 | Weak | Negative |
| apparent_temperature_max (°C) | 150 | 0.033 | Very weak / no correlation | Positive |

The most pronounced correlations were all negative and concentrated in variables related to humidity and precipitation: minimum, mean, and maximum relative humidity (*ρ* = *−*0.752, *−*0.749, *−*0.746, respectively), hours of precipitation (*ρ* = *−*0.728), and the total rain/precipitation (*ρ* = *−*0.703 for both—importantly, these two variables exhibited the same coefficient, indicating they could be redundant or nearly identical measures in the source data and should be examined for duplication prior to any subsequent multivariable modeling, such as regression, to prevent multicollinearity). This pattern indicates that case counts rise sharply as the atmosphere becomes drier.

A further set of comparably strong negative correlations supports this perspective: wet-bulb temperature (min, mean, max: *ρ* = *−*0.697, *−*0.660, *−*0.618), soil moisture (*ρ* = *−*0.626), daylight length (*ρ* = *−*0.612), and cloud cover (mean, min, max: *ρ* = *−*0.592, *−*0.587, *−*0.523) all displayed negative correlations with case counts, while maximum air temperature emerged as the most prominent positive correlate (*ρ* = 0.633). Wind direction exhibited a comparably sized negative correlation (*ρ* = *−*0.562), but as this variable is circular, the linear/monotonic interpretation of *ρ* must be approached with caution.

A more moderate tier of associations consisted of a combination of effects: minimum apparent temperature (*ρ* = *−*0.496) and surface pressure (minimum and average: *ρ* = *−*0.311, *−*0.304) exhibited negative correlations, while mean air temperature (*ρ* = 0.414) and mean soil temperature (*ρ* = 0.300) showed positive correlations. The weakest correlations were primarily driven by wind-related factors (maximum wind speed, average and maximum wind gusts: *ρ* = 0.289, 0.274, 0.269, all positive) as well as slight negative correlations for average apparent temperature, minimum temperature, and maximum surface pressure.

Taken together, the overall direction of effects is internally consistent: variables indexing drier, cooler-humidity, less cloudy, shorter-daylight conditions (low relative humidity, low precipitation, low cloud cover, low wet-bulb temperature, shorter daylight duration) are associated with higher case counts, while higher air and soil temperature are associated with higher case counts as well, a pattern broadly consistent with the dry-season transmission peak reported for Lassa fever in West African settings, where reduced humidity and rainfall coincide with increased rodent–human contact (e.g., through bush-burning-driven rodent migration into dwellings) and with greater environmental stability of aerosolized virus under low-humidity conditions [40, 41]. It is worth noting, however, that not all of the published literature agrees on the precise mechanism: at least one ecological study has instead linked higher rainy-season humidity to improved viral survival outside the host and increased rodent movement for breeding [27], so the directionality found here, while consistent with the majority dry-season-peak literature, should be discussed as one plausible mechanism among others rather than a settled causal pathway, especially since Spearman’s *ρ* establishes association, not causation.

### 4.3 Multi-Layer Cartographic Interpretation of Lassa Fever Hotspots

To interpret the occurrence of Lassa fever, the cartographic analysis was carried out in thematic clusters. First, the annual confirmed-case maps are presented in order to identify the states that repeatedly fall into the highest burden category. Then, the fixed contexts, namely land use, lithology, and elevation are interpreted in relation to these hotspot states. Finally, the annual rainfall maps are examined to assess whether the hotspot situations coincide with specific rainfall zones or whether rainfall should be interpreted mainly through seasonal rather than annual patterns. This structure allows the analysis to move from the epidemiological pattern to the environmental and sociogeographic context that may help explain the occurrence of increased confirmed cases. In Figures 5–7 the cartographic elaboration is based on NCDC surveillance data retrieved from [7, 8].

**Figure 5:**
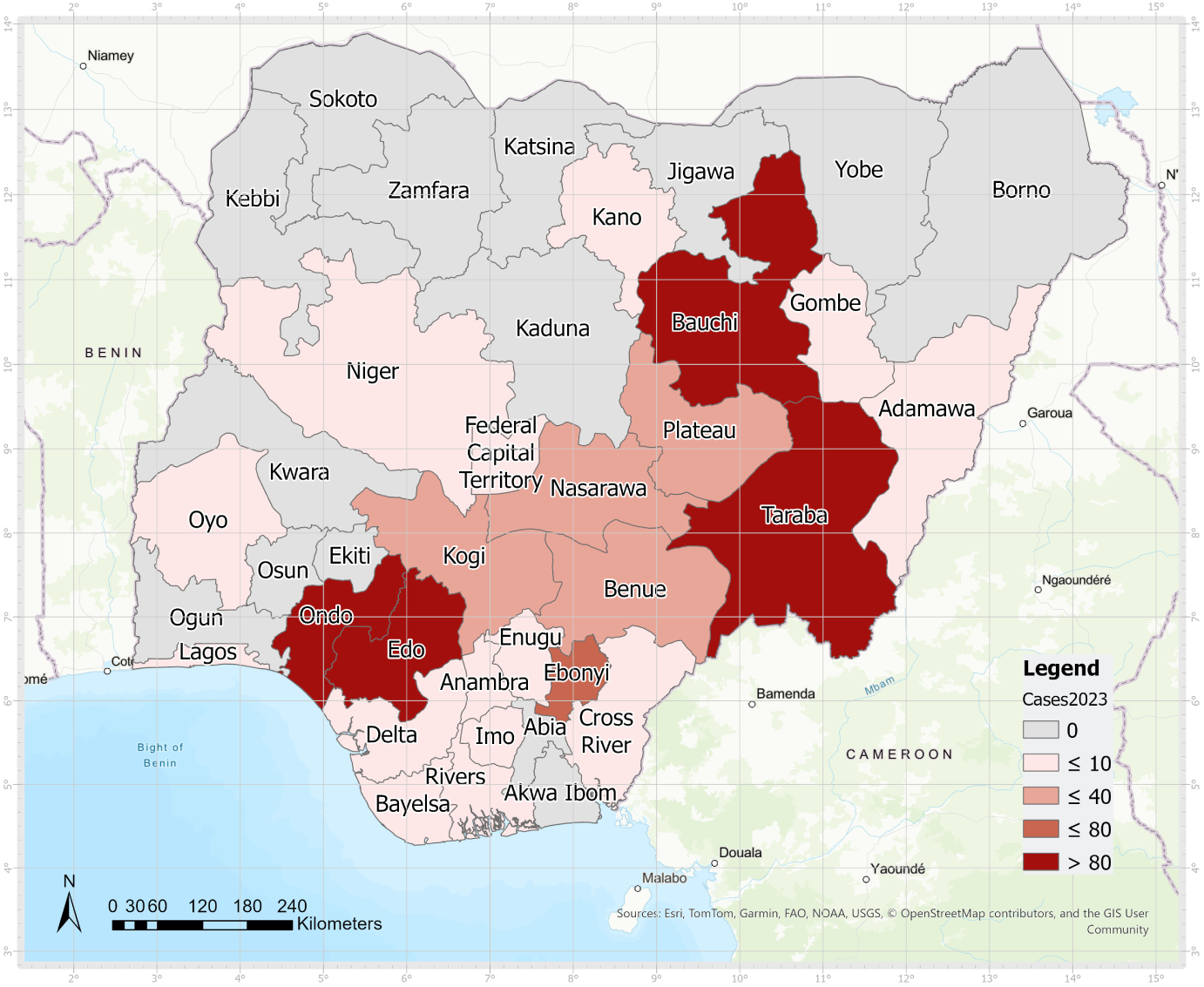
State-level distribution of confirmed Lassa fever cases in Nigeria in 2023.

**Figure 6:**
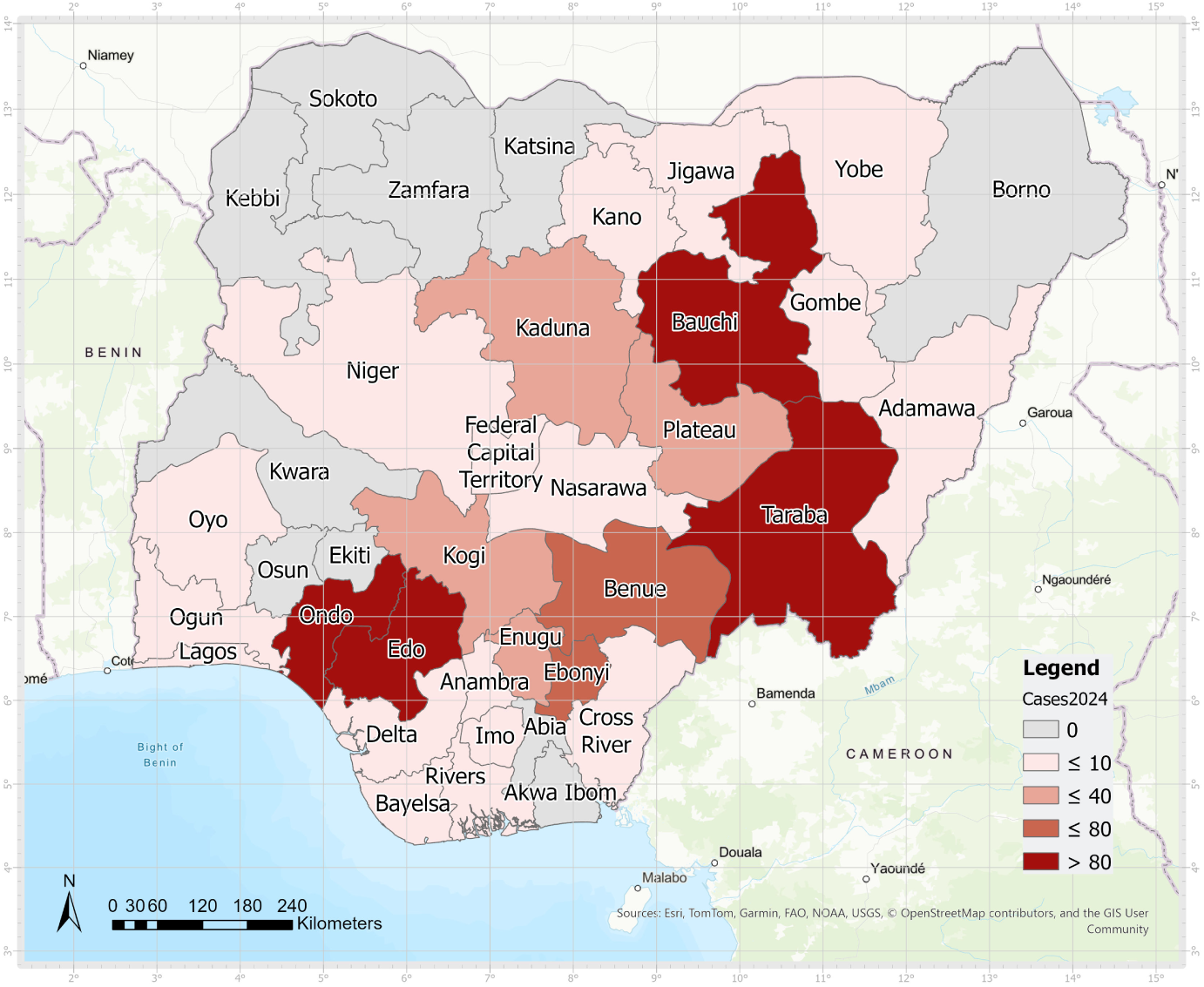
State-level distribution of confirmed Lassa fever cases in Nigeria in 2024.

**Figure 7:**
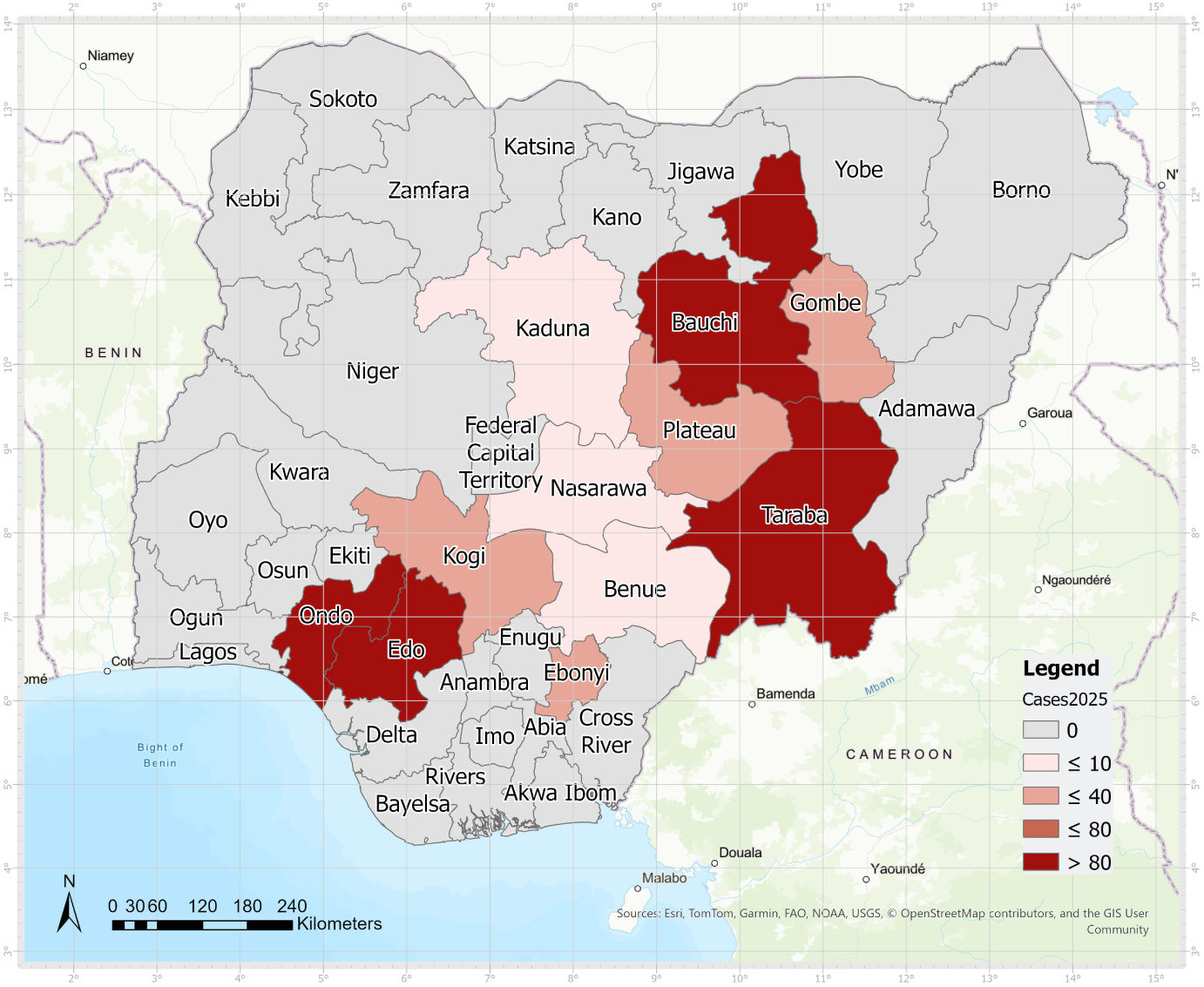
State-level distribution of confirmed Lassa fever cases in Nigeria in 2025.

#### 4.3.1 Annual Spatial Distribution of Confirmed Cases

The spatial core of the reported Lassa fever burden remains remarkably stable throughout the three-year study period, as shown in Figures 5–7. In every year, the same four states are assigned to the highest common cartographic class of more than 80 confirmed cases: **Ondo** and **Edo** in the southwest, and **Bauchi** and **Taraba** along the inland central-eastern axis. Two broader concentrations of cases can also be distinguished: a southwestern and southern cluster centred on Ondo and Edo and extending toward Ebonyi, and an inland central-eastern cluster involving Bauchi, Taraba, Plateau, Gombe, Nasarawa, Kogi and Benue. These two concentrations are connected by a zone of low-to-moderate burden across Plateau, Nasarawa, Kogi and Benue. This connecting pattern is relatively evident in 2023 and 2024, but becomes weaker and less spatially continuous in 2025.

Although the same four hotspot states remain within the highest class throughout the study period, the magnitude of the maximum state-level burden changes between years. As shown in Table 6, the highest state-level total was recorded in Ondo in all three years, reaching 358 cases in 2023, 359 cases in 2024 and 190 cases in 2025. These values are reported separately from the common open-ended cartographic class because all values above 80 cases are represented by the same colour in the annual maps. The table therefore retains information on the changing intensity of the most severely affected state, while the common map intervals preserve direct spatial comparability among years.

**Table 6:** Annual maximum number of confirmed Lassa fever cases recorded at the state level during the study period.

| Year | Maximum Number of Cases | State |
| --- | --- | --- |
| 2023 | 358 | Ondo |
| 2024 | 359 | Ondo |
| 2025 | 190 | Ondo |

The geographic contraction of the reported case distribution did not occur uniformly, but developed in two distinguishable phases. Between 2023 and 2024, several southern states that had recorded low case totals in 2023, including Delta, Imo, Cross River and Bayelsa, moved into the zero-case category, while the four core hotspot states and much of the inland connecting zone remained comparatively stable. A second and more pronounced contraction occurred between 2024 and 2025. During this phase, the burden declined across several north-central connecting states, particularly Kogi, Benue and Nasarawa, while Ebonyi also shifted toward a lower case category compared with its higher burden in 2023. Consequently, the 2025 map shows a more spatially concentrated distribution, with fewer states reporting confirmed cases and a clearer concentration around the persistent southwestern and central-eastern hotspot axes.

The decline in the annual maximum from 359 cases in 2024 to 190 cases in 2025 indicates a substantial reduction in the intensity of the highest state-level burden. However, the continued presence of Ondo, Edo, Bauchi and Taraba in the highest common class demonstrates that the geographical structure of the principal hotspots remained stable despite the overall contraction. Thus, the 2025 pattern reflects a reduction in the magnitude and spatial spread of reported transmission rather than a complete reorganization of the established hotspot geography.

#### 4.3.2 Land-Use Context of Hotspot States

Figure 8 illustrates the land-use settings of the hotspot states shown in Figures 5–7. The extremely high-burden states are not confined to just one type of land cover. Ondo and Edo, found in the southwestern hotspot, are placed in a moist southern setting where forested regions, farmland, and urban areas exist in close spatial relation. This implies that the hotspot is situated within a mosaic environment instead of being solely in an urban, agricultural, or forested area.

**Figure 8:**
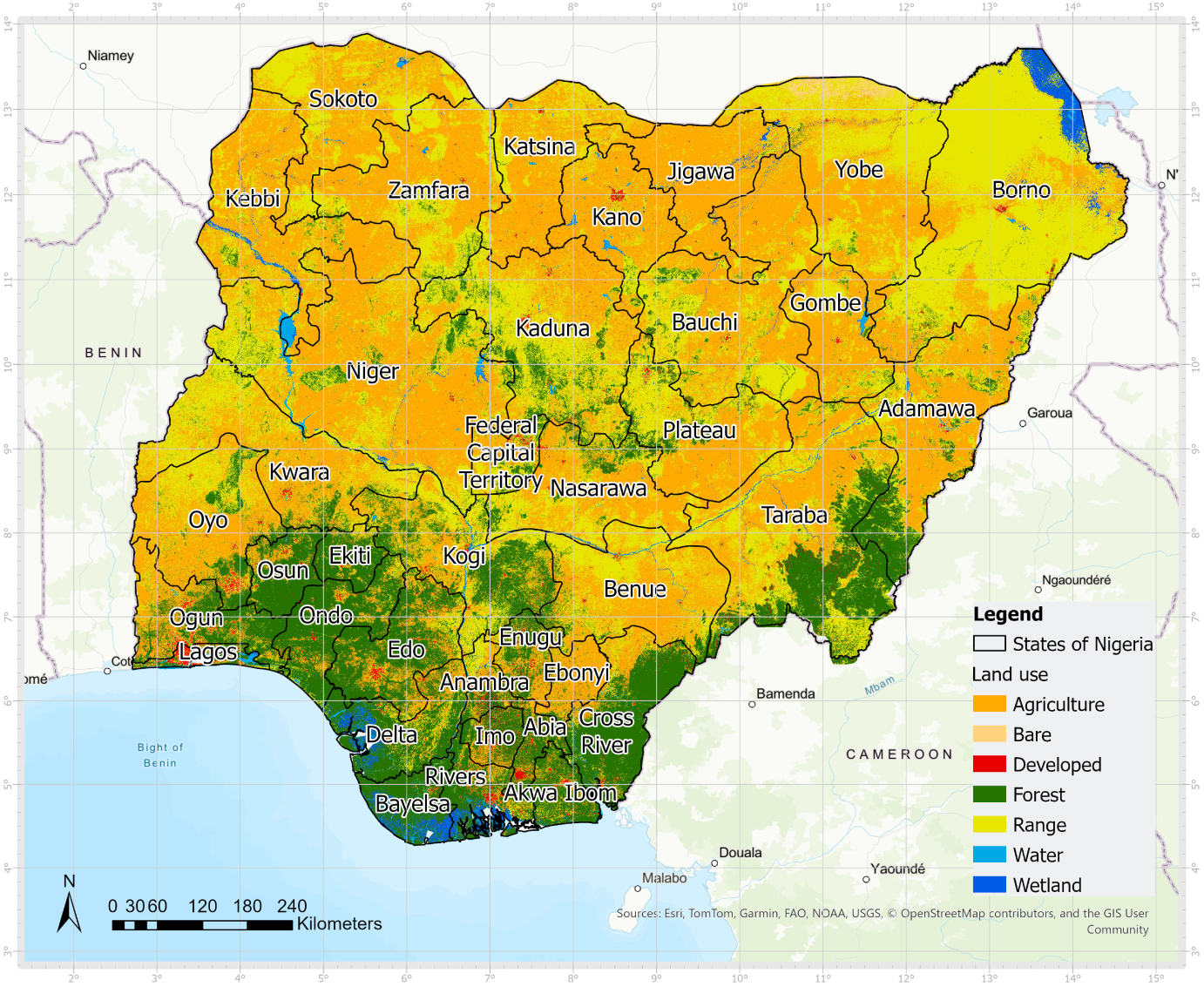
Land-use map of Nigeria. Source: authors’ cartographic elaboration using the study GIS layers [13]

Such mixed landscapes are epidemiologically relevant because they combine human settlement, farming activity and food storage in close proximity to the rodent reservoir *Mastomys natalensis*, whose occupancy has been shown to increase along a gradient from forest toward agricultural and village habitats, with the species essentially absent from undisturbed forest itself [42]. Agricultural production and household food storage practices are recognized drivers of rodent population dynamics and can attract the reservoir close to human activity [43], while remaining forested or semi-natural patches embedded within the mosaic may still provide cover and movement corridors connecting agricultural plots, even if they are not the primary habitat of the reservoir itself. Therefore, in Ondo and Edo, the land-use pattern supports the interpretation of repeated rodent–human contact opportunities concentrated at the agriculture–settlement interface of the forest–agriculture–settlement mosaic, consistent with land-use-gradient studies elsewhere in West Africa [42, 43].

In contrast, Bauchi and Taraba are located further inland and are associated with more transitional land-use conditions. Bauchi is located in a zone where agricultural and pastoral environments are more prominent, while Taraba contains a heterogeneous combination of agricultural land, range areas and forested or semi-natural zones. This contrast suggests that very high Lassa fever incidence is not restricted to one ecological setting. Rather, the hotspot states possess a common trait: they exist in regions where human actions intersect with natural or semi-natural surroundings. This supports the interpretation that land-use complexity may contribute to repeated opportunities for rodent–human contact, especially where farming, household food storage and settlement expansion interact [43].

#### 4.3.3 Lithological Background and Environmental Setting

Figure 9 provides the lithological context of Nigeria. The purpose of including this level is not to suggest a direct causal relationship between rock type and the occurrence of Lassa fever, but to examine the physical substrate that may influence drainage, soil development, moisture retention, geomorphology, and the distribution of land use patterns. These factors are relevant because they can indirectly influence the environmental conditions under which rodent habitats and human activities may overlap. In the legend of the lithological map, the class **N/A** refers to areas for which lithological information was *not available* in the source dataset; therefore, these areas were not interpreted as a separate lithological category.

**Figure 9:**
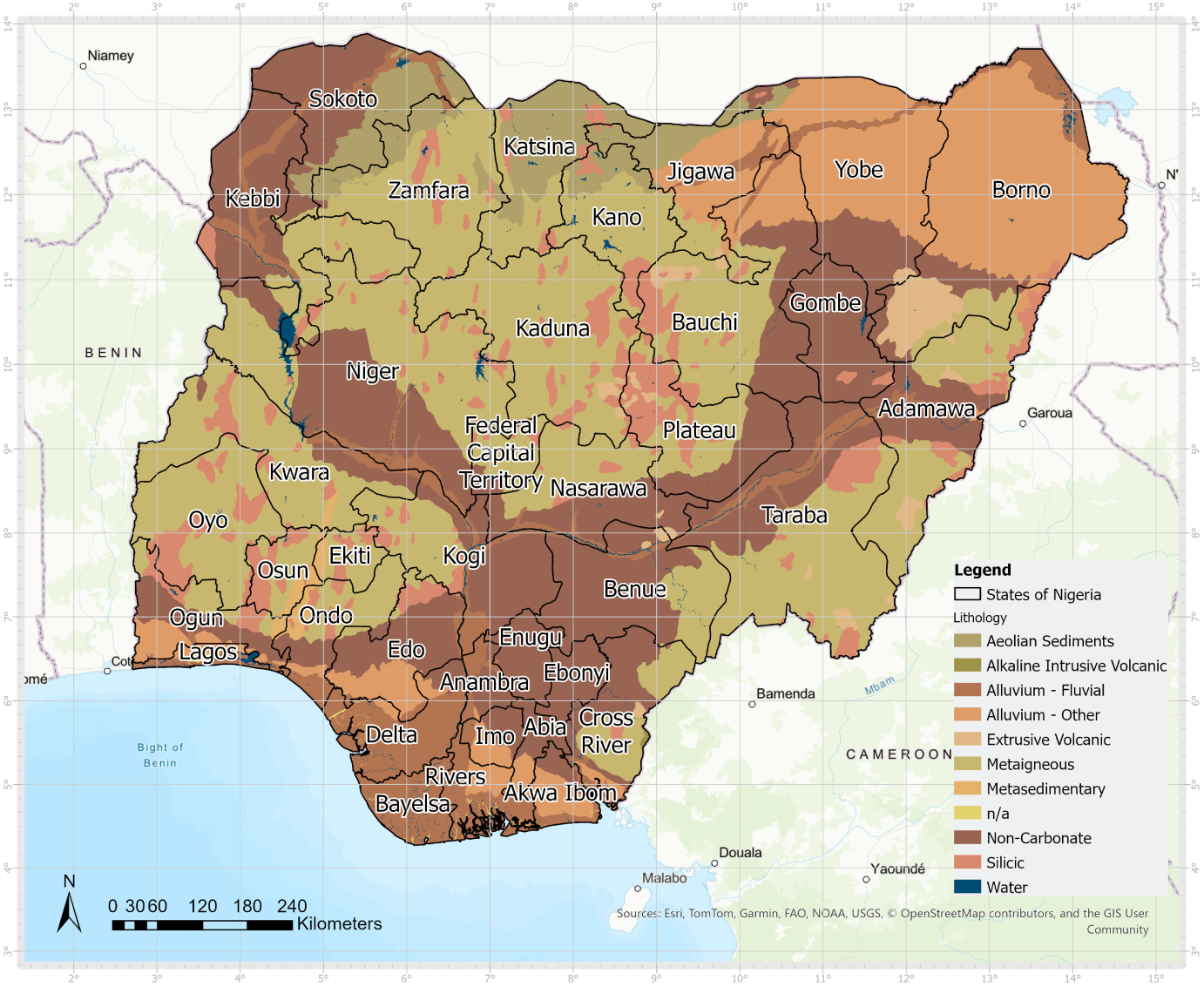
Lithological map of Nigeria based on data from Regional Centre for Mapping of Resources for Development, Source: authors’ cartographic elaboration based on RCMRD/USGS data [12]

In the southwestern area, Edo and Ondo are located near alluvial/fluvial deposits, non-carbonate formations, metasedimentary units, and siliceous formations. The existence of alluvial and fluvial units is especially significant as these deposits are often linked to lowland zones, river pathways, and regions with increased soil moisture and agricultural productivity. The combination of these lithological conditions with the forest-agricultural mosaic shown in Figure 8 presents a situation where cropland, human settlements, food storage practices, and rodent habitats can coexist [43].

In Bauchi and Taraba, the geological environment is more varied, consisting of metasedimentary, non-carbonate, alluvial, and at times volcanic or silicic materials. This variety could influence irregular changes in drainage, soil characteristics, vegetation composition, and land utilization. Such scenarios also occur in transitional inland regions, where agriculture, grazing lands, and semi-natural habitats exist together. The analysis of Figures 8 and 9 indicates that the inland hotspot is influenced by various lithological units, resulting from the interplay of natural substrate, land use, hydrological elements, and human impact.

#### 4.3.4 Elevation and Geomorphological Context

Figure 10 adds a geomorphological perspective to the interpretation of the hotspot states. Elevation is not considered a direct causal factor of Lassa fever occurrence; however, it can influence several envi-ronmental conditions that are relevant to rodent ecology and human exposure, including temperature, soil moisture, drainage, vegetation structure, agricultural suitability and settlement distribution. For this reason, elevation is used here as a contextual layer that helps describe the physical setting within which land use, rainfall seasonality and rodent–human interaction may operate.

**Figure 10:**
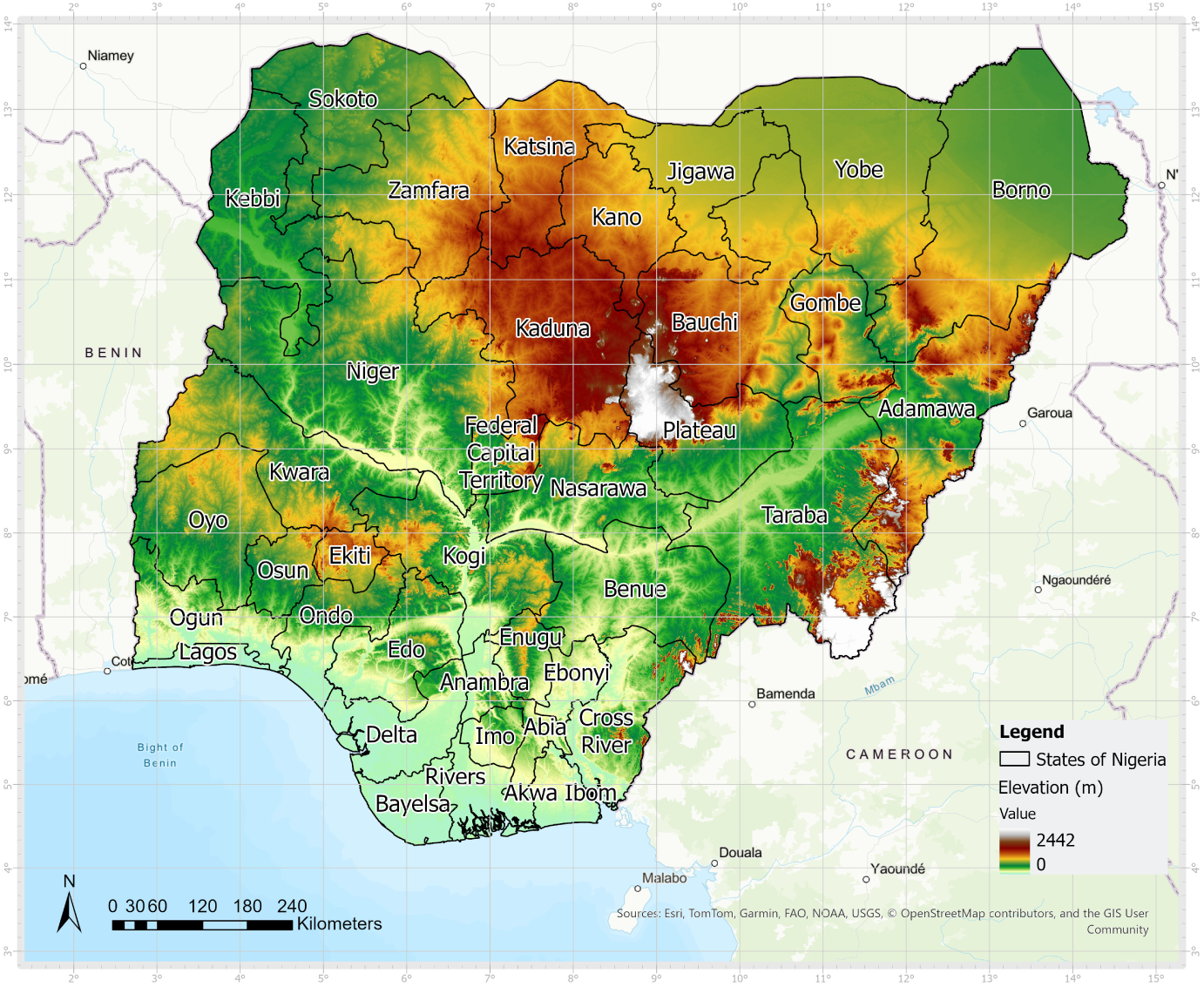
Elevation map of Nigeria, Source: authors’ cartographic elaboration based on NASA - EARTHDATA data [11].

The southwestern hotspot states, Ondo and Edo, are mainly located in lower-elevation southern environments. These areas coincide with humid conditions, forest–agriculture mosaics and lithological settings that include alluvial/fluvial and non-carbonate formations. Such lowland and humid environments can support agricultural activity, food storage, human settlement and vegetation cover, all of which may increase opportunities for the rodent reservoir to occur near human activity, particularly given its association with agricultural and village habitats rather than undisturbed forest [42, 43]. In this sense, elevation contributes to the interpretation of Ondo and Edo as lowland hotspot environments where human settlement, farming and suitable rodent-supporting habitats may coexist.

By contrast, Bauchi and Taraba are located in more inland and topographically variable settings. These areas include higher or more heterogeneous elevation zones compared with the southwestern states. This geomorphological variability may influence drainage, local moisture availability, vegetation patterns and land-use transitions. In combination with the lithological heterogeneity shown in Figure 9 and the mixed land-use structure shown in Figure 8, elevation helps explain why the inland hotspot states may represent different but still suitable environmental contexts for repeated rodent–human interaction.

#### 4.3.5 Annual Precipitation Context

Figures 11–13 provide the annual precipitation context for the hotspot states. Throughout the years, the maps illustrate the extensive north-south rainfall gradient in Nigeria, showing wetter areas in the southern and southeastern regions, with diminishing rainfall amounts as one moves northward. Amid this gradient, the high-burden states are situated in diverse rainfall zones. Ondo and Edo are positioned within the moist southern region, whereas Bauchi relates to a more moderate rainfall environment; Taraba, by contrast, spans an internal gradient, with its north-western lowlands falling in the moderate range while its south-eastern highlands (the Mambilla Plateau area) receive rainfall in excess of 2,000 mm, comparable to the southern hotspot states.

**Figure 11:**
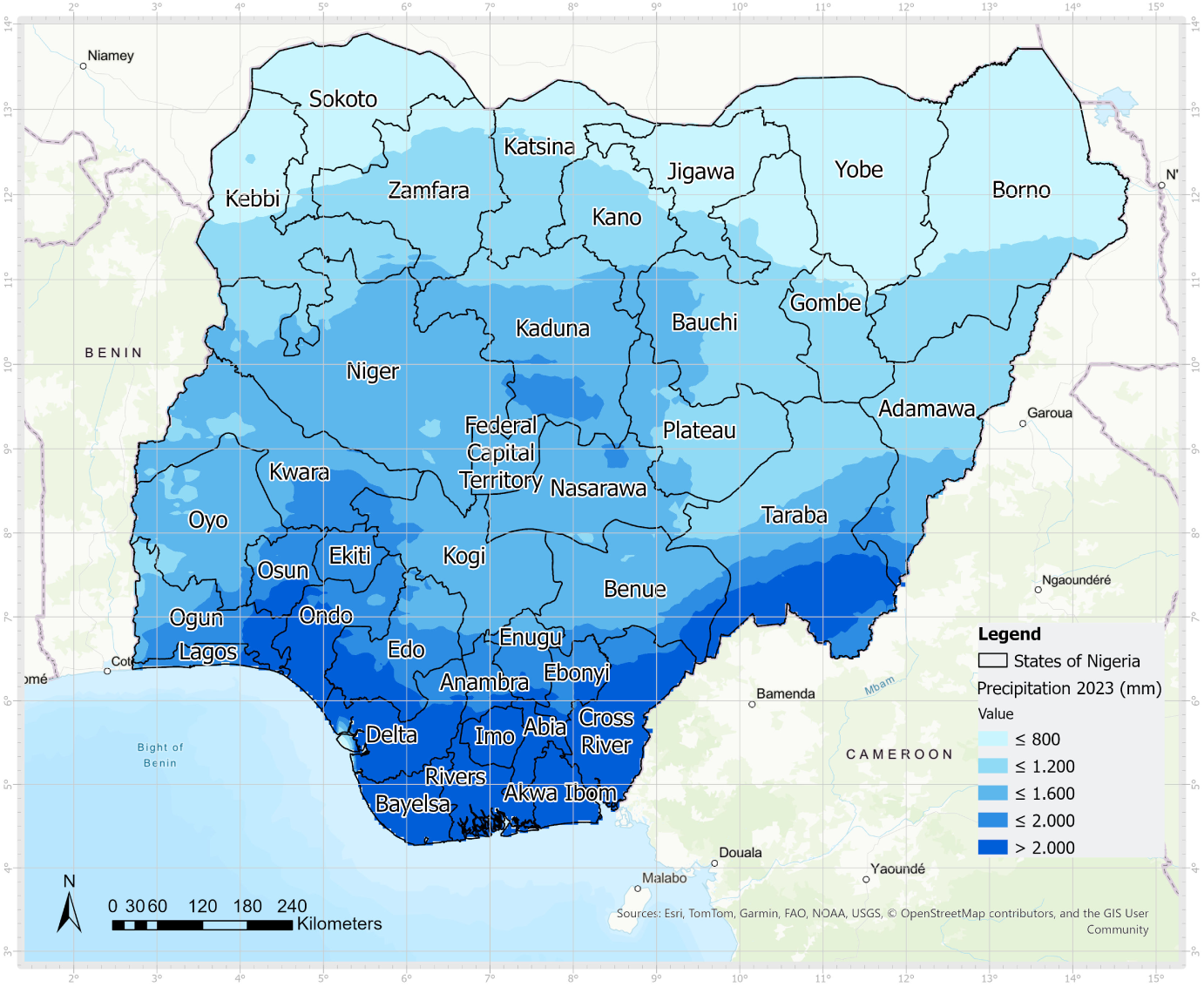
Annual precipitation in Nigeria in 2023. Source: authors’ cartographic elaboration based on CHIRPS rainfall data [10].

**Figure 12:**
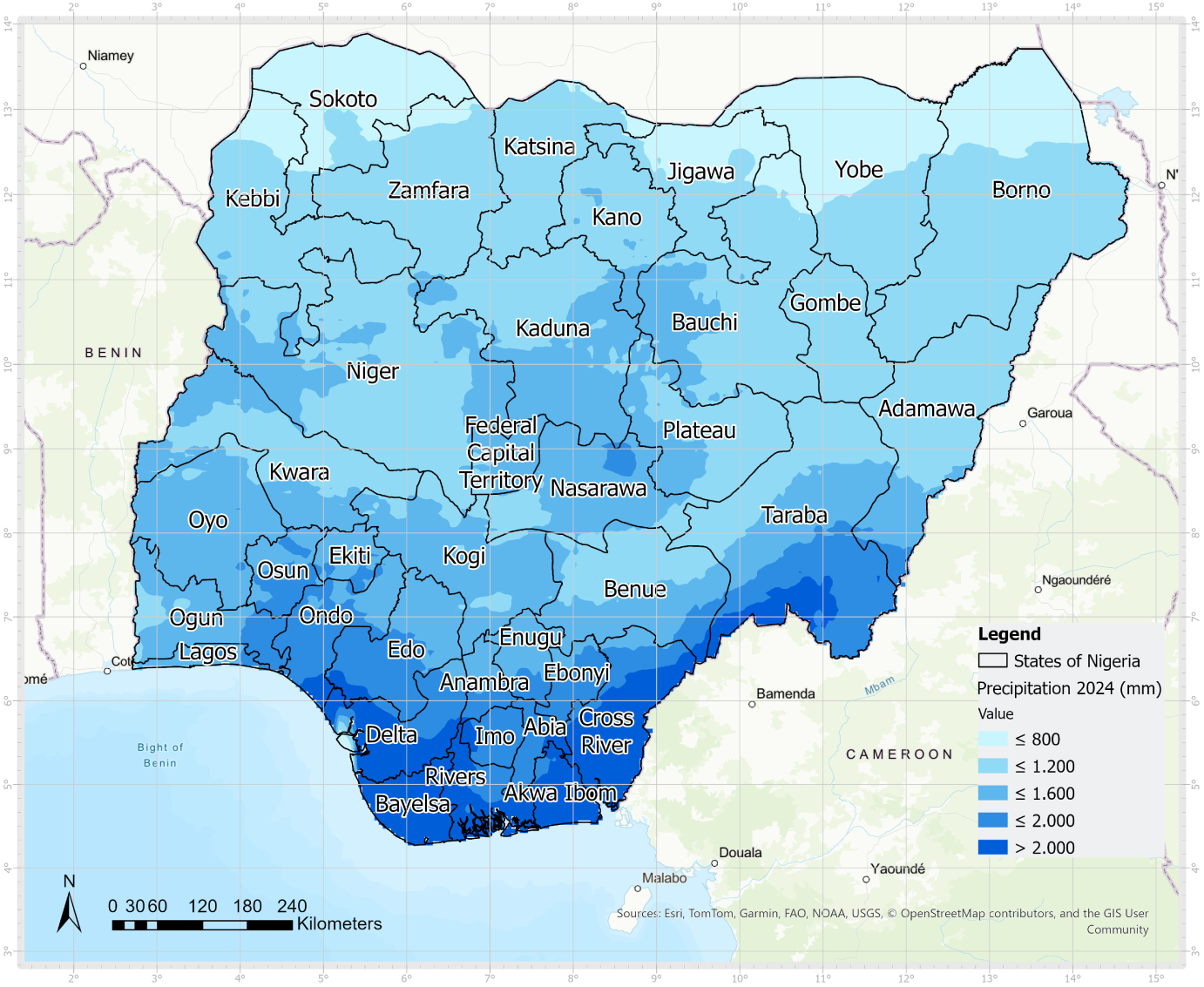
Annual precipitation in Nigeria in 2024. Source: authors’ cartographic elaboration based on CHIRPS rainfall data [10].

**Figure 13:**
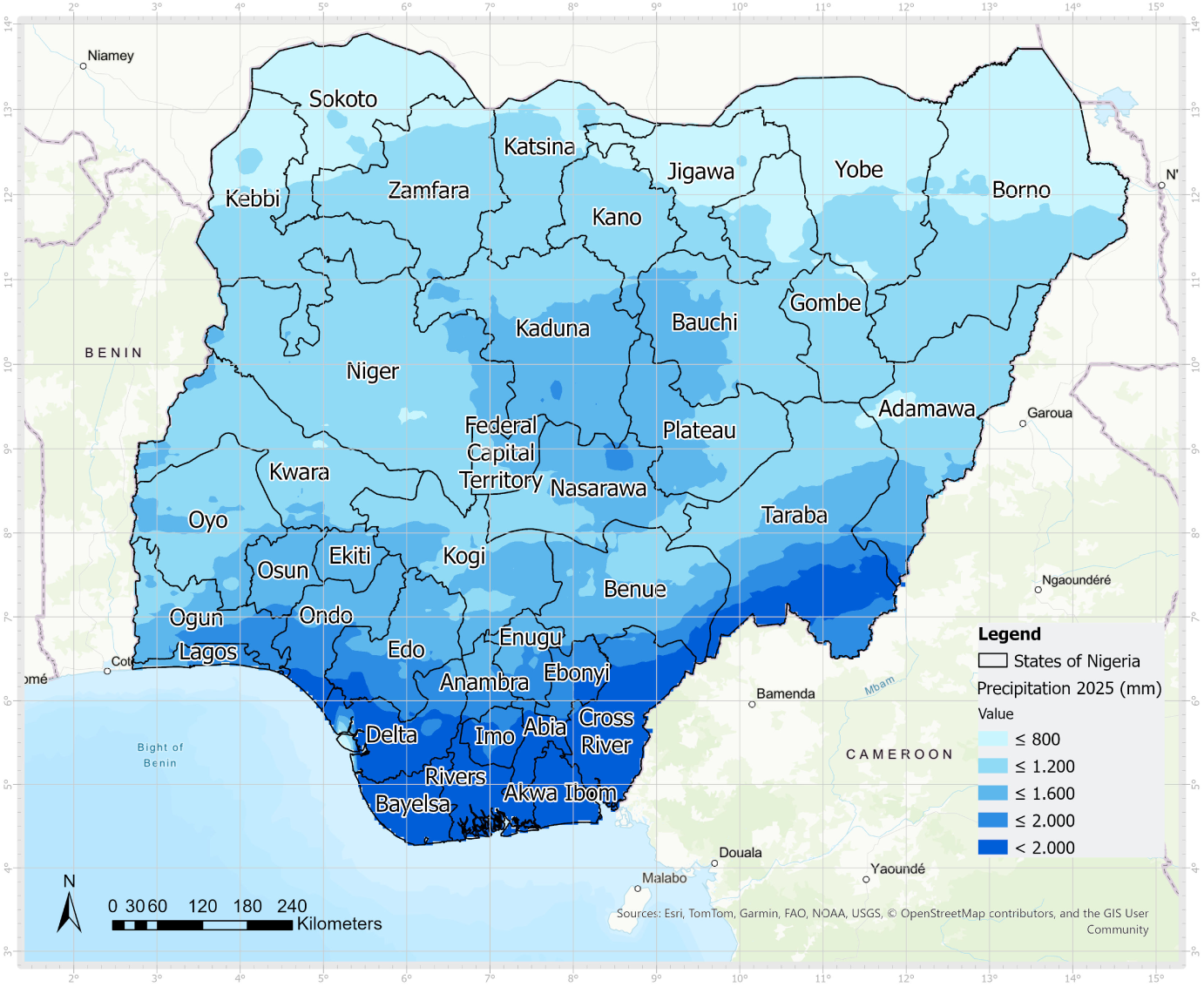
Annual precipitation in Nigeria in 2025. Source: authors’ cartographic elaboration based on CHIRPS rainfall data [10].

The 2023 rainfall map indicates that the southwestern hotspot states align with the southern wetter rainfall region, whereas Bauchi and the lowland parts of Taraba are situated in more transitional inland rainfall areas. The 2024 map verifies a comparable structure, showing that the overall precipitation trend stays consistent and that the same hotspot states continue to exist even though they belong to various rainfall zones. The 2025 map further supports this interpretation: although the annual maximum state-level case count changes, the hotspot geography remains similar under the same broad precipitation gradient.

This pattern suggests that high annual rainfall totals alone do not fully explain the spatial distribution of Lassa fever. In conjunction with the diagrams in the previous section, we agree that time and seasons must be taken into account. Moreover, the recurrence of the same hotspot states suggests that rainfall-related mechanisms are more likely connected to seasonal transitions, dry-season exposure windows, vegetation cycles, rodent food resources and household storage practices rather than to annual totals alone [43]. Therefore, annual precipitation maps provide an important environmental background, but future spatial models should include precipitation variables at a finer temporal scale, especially weekly or seasonal rainfall indicators.

#### 4.3.6 Synthesis of the Multi-Layer Interpretation

The comprehensive cartographic examination indicates that the major impact of Lassa fever is consistently focused in Ondo, Edo, Bauchi, and Taraba. These areas possess unique environmental features. Nonetheless, they are found in areas where differences in climate, land use patterns, geological characteristics, elevation, water movement, and human-environment interactions can converge to form ongoing threats. The joint evaluation of the reference layers indicates that Lassa fever hotspots should not merely be viewed as administrative regions with high case counts, but rather as socio-ecological environments where rodent habitats, farming methods, food storage techniques, and seasonal climate variations might intersect [42, 43].

Land use highlights areas where agriculture, vegetation and urban areas intersect. Lithology indicates the geological base that may influence soil and drainage conditions. Elevation offers information on soil and microclimatic differences. Precipitation outlines the hydroclimatic environment. Collectively, these layers facilitate a more coherent understanding of Lassa fever hotspots and highlight their importance in upcoming hotspot assessments, early warning systems, and focused public health efforts.

## 5 Discussion

The results support the interpretation that Lassa fever risk in Nigeria is strongly seasonal and spatially selective. The weekly analysis indicates that outbreak weeks are concentrated mainly in a dry-season window extending from approximately December to March, and that rainfall, precipitation hours and humidity provide important seasonal susceptibility signals [22, 40]. From a conventional epidemiological perspective, this finding contributes to the understanding of Lassa fever seasonality.

This pattern is quantitatively reinforced by the Spearman rank-correlation analysis performed between the 28 environmental parameters and weekly confirmed case counts (Table 5). The majority of the parameters examined showed comparatively large coefficients, with the strongest associations—all negative—observed for relative humidity (*ρ* ranging from *−*0.752 to *−*0.746), precipitation hours (*ρ* = *−*0.728), and rainfall/precipitation sum (*ρ* = *−*0.703). Maximum air temperature was the strongest positive correlate (*ρ* = 0.633), and a comparable negative pattern was observed for wet-bulb temperature, soil moisture, daylight duration and cloud cover. Only maximum apparent temperature showed a coefficient close to zero (*ρ* = 0.033), suggesting no substantial monotonic relationship with case occurrence. Taken together, these coefficients indicate that case counts rise sharply as conditions become drier, less humid and less cloudy, and as air and soil temperature increase—consistent with the dry-season transmission peak reported elsewhere for Lassa fever in West Africa [22, 40].

The cartographic analysis strengthens this interpretation in several ways. Initially, the yearly case maps show ongoing spatial concentration in a limited number of states, particularly Ondo, Edo, Bauchi, and Taraba. This persistence indicates that the distribution of Lassa fever in Nigeria is organized instead of arbitrary. Secondly, the land-use map shows that these hotspot states are situated within varied environmental contexts where agricultural practices, forests or transitional vegetation, and human habitation coincide. Such landscapes may favor repeated rodent–human contact, especially where food storage, housing quality and sanitation are inadequate. Finally, the CHIRPS precipitation maps demonstrate that the relevant signal lies not in annual rainfall totals by themselves, but in the seasonal organization of wet and dry periods within those broader climatic environments.

A key methodological contribution of the study is the transformation of heterogeneous datasets into a coherent spatial decision-support structure. Surveillance time series, weekly weather indicators, annual precipitation rasters and land-use classes were not treated as separate products, but as complementary levels of interpretation; the Spearman correlation analysis served as the quantitative bridge linking the weekly weather indicators to case incidence before this information was carried into the spatial layers.

The relationship with smart-sensor and AI-based approaches is also important. The ongoing study relies on accessible surveillance and gridded climate data; nonetheless, this approach could be improved by integrating near-real-time rainfall gauges, river-stage monitoring, soil-moisture sensors, radar rainfall information, drone-gathered observations, and field assessments. These data feeds might supply machine-learning models designed to assess vulnerability under varying circumstances. In this regard, the existing workflow serves as an initial move toward a dynamic vulnerability platform: it currently lacks real-time sensing or predictive modeling, yet it lays the groundwork for the data logic, spatial integration, and cartographic communication framework necessary for this strategy.

A plausible ecological interpretation is that the rainy season influences the reproductive cycle, habitat use and food-search behavior of the rodent reservoir, and that the subsequent dry period brings infected rodents into closer contact with human habitations [22, 40]. This is consistent with the negative association between humidity/rainfall and case counts and the positive association with temperature identified in the correlation analysis (Table 5), although these bivariate coefficients establish association rather than causation, and several predictors (e.g., rainfall and precipitation sum, which returned identical coefficients) were highly collinear and should be interpreted jointly rather than as independent signals. Under this interpretation, Lassa fever outbreaks represent the outcome of a coupled socioecological system in which climate, land use, household practices and health-system responsiveness interact. The maps do not prove this mechanism directly, but they are compatible with it and provide a strong exploratory basis for more formal spatial modelling.

From a public health perspective, the most valuable finding is the existence of a location-specific and seasonally stable high-risk period. If higher-risk weeks mainly occur from December to March, the spread of awareness messages, the execution of rodent control measures, food storage advice, and healthcare preparedness can be increased before and during this timeframe. As the hotspot states recur annually, focused actions in Ondo, Edo, Bauchi, and Taraba would probably be more effective than a nationwide approach.

## 6 Conclusions

This manuscript presents a pilot application of multi-layer GIS-based susceptibility mapping for Lassa fever and geographic information. The Lassa fever analysis for Nigeria during 2023–2025 indicates a recurring seasonal pattern in which confirmed cases increase during dry weeks with near-zero rainfall and decline during wetter periods. Rainfall, precipitation hours and humidity were the most informative weather indicators in the descriptive analysis, returning the strongest Spearman rank correlations with weekly case counts among the 28 environmental parameters examined, with coefficients reaching *ρ* = *−*0.75 in magnitude; air temperature was the strongest positive correlate (*ρ* = 0.633).

The integration of thematic maps significantly strengthens the interpretation. Ondo, Edo, Bauchi and Taraba emerged as persistent hotspot states across multiple years, while the land-use and CHIRPS precipitation maps helped show that hotspot persistence cannot be explained by a single factor alone. Instead, the evidence points to a combined role of seasonal rainfall dynamics, mixed environmental settings and local human–environment interaction.

The main contribution of the paper, as stated earlier, is methodological. Initially, it demonstrates how varied datasets can be collected and analyzed with Python, subsequently evaluated statistically using rank-correlation analysis, and ultimately visualized via maps. Future studies should thus broaden the current workflow by integrating radar and field data, high-resolution remote sensing, spatial databases, and modeling, leveraging the environmental predictors found here to be most closely linked with case incidence. This expansion would facilitate the development of dynamic vulnerability maps for local governments, disaster response systems, and resilient community planning.

## Data Availability

The datasets generated and analyzed during this study, including the Lassa fever case data by state and local government area (LGA), associated weather parameters, and derived outbreak-week classifications, are openly available in the GitHub repository in [44].

